# Negative Contextual Valence Unmasks Altered Counterfactual Decision-Making in Major Depressive Disorder

**DOI:** 10.64898/2026.05.15.26353249

**Authors:** Avijit Chowdhury, Philipp Neukam, Ofer Perl, Matthew Heflin, Jacob Yael, Laurel S. Morris, Xiaosi Gu, James W. Murrough

**Affiliations:** The Dennis S. Charney, MD, Depression and Anxiety Discovery Center, Icahn School of Medicine at Mount Sinai, New York, NY, United States; Center for Computational Psychiatry, Icahn School of Medicine at Mount Sinai, New York, NY, United States; Department of Cognitive Sciences, University of Haifa, Israel; Department of Psychiatry, Yale School of Medicine, New Haven, CT, United States; Department of Experimental Psychology, University of Oxford, Oxford, United Kingdom; Nuffield Department of Clinical Neurosciences, University of Oxford, Oxford, United Kingdom; Department of Biomedical Informatics and Data Science, Yale School of Medicine, New Haven, CT, United States; Nash Family Department of Neuroscience, Icahn School of Medicine at Mount Sinai, New York, NY, United States; VISN 2 Mental Illness Research, Education and Clinical Center (MIRECC), James J. Peters VA Medical Center, Bronx, NY, United States

**Keywords:** Depression, Regret, Anhedonia, Neuroimaging, Fictive error

## Abstract

**Background:** While counterfactual thinking (’what could have been’) guides adaptive decision-making, it remains unclear how this process is altered by the negative biases and motivational deficits characteristic of Major Depressive Disorder (MDD).

**Methods:** We used a sequential economic decision-making task designed to emulate a volatile stock market to assess choice behavior in adults with or without MDD (Total N=178); a subset of these participants completed the task during functional MRI (N=53). The task allowed participants to make either positive (’invest’) or negative (’short’) bets, under either positive or negative contextual valence—defined by whether the immediately preceding stock price change was positive or negative. Fictive errors were defined as the difference between realized and best-possible outcomes.

**Results:** Across the full cohort, group differences in behavioral adjustments to fictive error signals emerged exclusively under negative contextual valence, when stock prices decreased. Compared with controls, participants with MDD showed heightened sensitivity to invest-and-loss fictive errors, reflected in a greater reduction in subsequent bets (interaction β = −0.63, p < .001), but blunted adjustment to short-and-gain fictive errors (β = −0.86, p < .001). In the imaging cohort, blunted short-and-gain adjustment was accompanied by heightened anterior cingulate (ACC) activity and attenuated ventromedial prefrontal (vmPFC)-to-ACC coupling in MDD. vmPFC activity following negative market returns also tracked depression symptom severity.

**Conclusions:** Depression selectively disrupts the use of counterfactual outcomes to guide adaptive choice under negative contextual valence, implicating altered frontocingulate function in maladaptive decision-making.

**Significance Statement:** Counterfactual thinking about what might have happened contributes to adaptive decision-making, yet how it is altered in depression has remained unclear. Combining behavioral and functional MRI data, we show that major depressive disorder is not associated with a global impairment in counterfactual learning. Instead, deficits emerged selectively following negative market movements: depressed individuals over-adjusted after losses that confirmed the negative setting but failed to adapt after gains that contradicted it. These behavioral abnormalities were accompanied by altered anterior cingulate and ventromedial prefrontal function. The findings identify a context-dependent mechanism by which depression distorts the use of counterfactual outcomes to guide future choice.

## Introduction

A critical aspect of adaptive behavior is the ability to learn not only from experienced outcomes, but also from counterfactual outcomes – what could have occurred had a different action been taken (1–3). This capacity is formalized in computational models of behavior through two distinct signals: 1) (standard) experienced prediction errors, which arise from the discrepancy between actual and expected outcomes, and 2) fictive errors, which stem from the difference between the actual outcome and the unchosen alternative (4–7). Fictive error signals, sometimes interpreted as computational markers of regret (1), arise from a sense of instrumentality – the feeling that one’s choice produced the outcome – and become most pronounced when an alternative, better action was possible (8, 9). These signals often prompt “regret-averse” adjustments in behavior (5, 6, 10, 11). Aberrant neurobehavioral processing of fictive error signals has been associated with maladaptive decisionllJmaking in addiction (5) and in psychopathic individuals (12). Findings from these studies suggest that intact neural or subjective experience of counterfactual emotions does not always lead to adaptive behavioral adjustment, consistent with the idea that counterfactual information relies on additional evaluative processes to influence learning (13). However, it remains unclear how fictive error processing is altered in Major Depressive Disorder (MDD), a condition characterized by negative evaluative biases and impaired adaptive decision-making (14, 15).

Major depressive disorder involves disruptions in the cognitive and affective processes that support the evaluation and integration of action outcomes (15, 16). A core feature of depression is anhedonia, which includes impairments in motivation and reward-based learning (17, 18). MDD is also marked by a mood-congruent negativity bias (19) and heightened appraisal of negative events (20, 21). These features may be especially relevant under volatile action-outcome contingencies, where uncertainty may alter the sense that outcomes depend on one’s choices (22–24) and thereby disrupt evaluation of alternative actions. At the neural level, this is reflected in dysfunction across frontostriatal and frontocingulate circuits involved in reward appraisal, outcome monitoring, and evaluative control (25, 26). Collectively, this neurobehavioral profile underlies difficulties in learning and decision-making based on environmental feedback (15), processes typically formalized as reinforcement-learning (RL) (27).

While RL deficits are a hallmark of depression (15, 28), the mechanisms driving these impairments remain unclear. A common proposal is blunted reward sensitivity (29), with depressed individuals showing reduced reward bias and diminished learning from positive outcomes (30, 31). Findings regarding learning from losses are mixed, with some reporting excessive avoidance learning while others report no differences compared to controls (28, 29, 32, 33) – an inconsistency further confounded by comorbid anxiety (15, 34). These mixed findings in reward and loss learning may reflect that individuals respond not only to outcome valence, but also to how outcomes are evaluated relative to the best foregone alternative. Fictive errors capture this counterfactual comparison and are especially relevant for evaluating controllable actions rather than purely probabilistic gains and losses. Their influence depends on anticipated outcomes, decision context, and agency (10, 11, 13, 35, 36), and shapes whether a missed opportunity diminishes the value of a gain or whether the absence of a better alternative mitigates the impact of a loss. More broadly, outcomes may also be weighed against the valence of the surrounding decision context, consistent with evidence that depressive behavioral deficits emerge specifically in overall negative (“poor”) contexts (37).

Despite its relevance, how fictive errors shape decision-making in depression remains poorly understood. Some studies suggest blunted behavioral adaptation to regret signals (38), whereas others report heightened sensitivity to missed opportunities (39), consistent with reports of increased subjective regret (40). These mixed findings may reflect that counterfactual processing is not unitary, but engages multiple systems involved in valuation, agency, and behavioral control. Meta-analytic evidence suggests that these include ventral striatal, ventromedial prefrontal, and anterior cingulate regions (41–43). Because these circuits are also implicated in the pathophysiology of depression (16, 18, 44), they represent a plausible neural substrate for altered counterfactual processing.

We hypothesized that depression alters how fictive error signals influence subsequent behavioral adjustment, and that this effect is modulated by contextual valence. To test this, we combined functional MRI with a sequential economic decision-making task designed to emulate a volatile stock market, allowing us to assess counterfactual processing under positive and negative contextual valence (Figure 1). In this task, contextual valence refers to the positive or negative direction of the immediately preceding market return, irrespective of the participant’s choice. Participants could place positive bets by investing or negative bets by shorting, allowing gains and losses to occur under both positive and negative contextual valence. This design produced four outcome types: invest-gain, invest-loss, short-gain, and short-loss. We measured behavioral and neural responses to counterfactual, or fictive errors—defined as the difference between realized outcomes and foregone best alternatives—and tested their influence on subsequent choice. Group differences in fictive-error–driven behavioral change emerged only under negative contextual valence: depressed individuals over-adjusted after invest-losses but showed reduced adjustment after successful short-gains. This was accompanied by altered vmPFC and ACC function, implicating frontocingulate dysfunction in maladaptive learning in depression.

**Figure 1.**
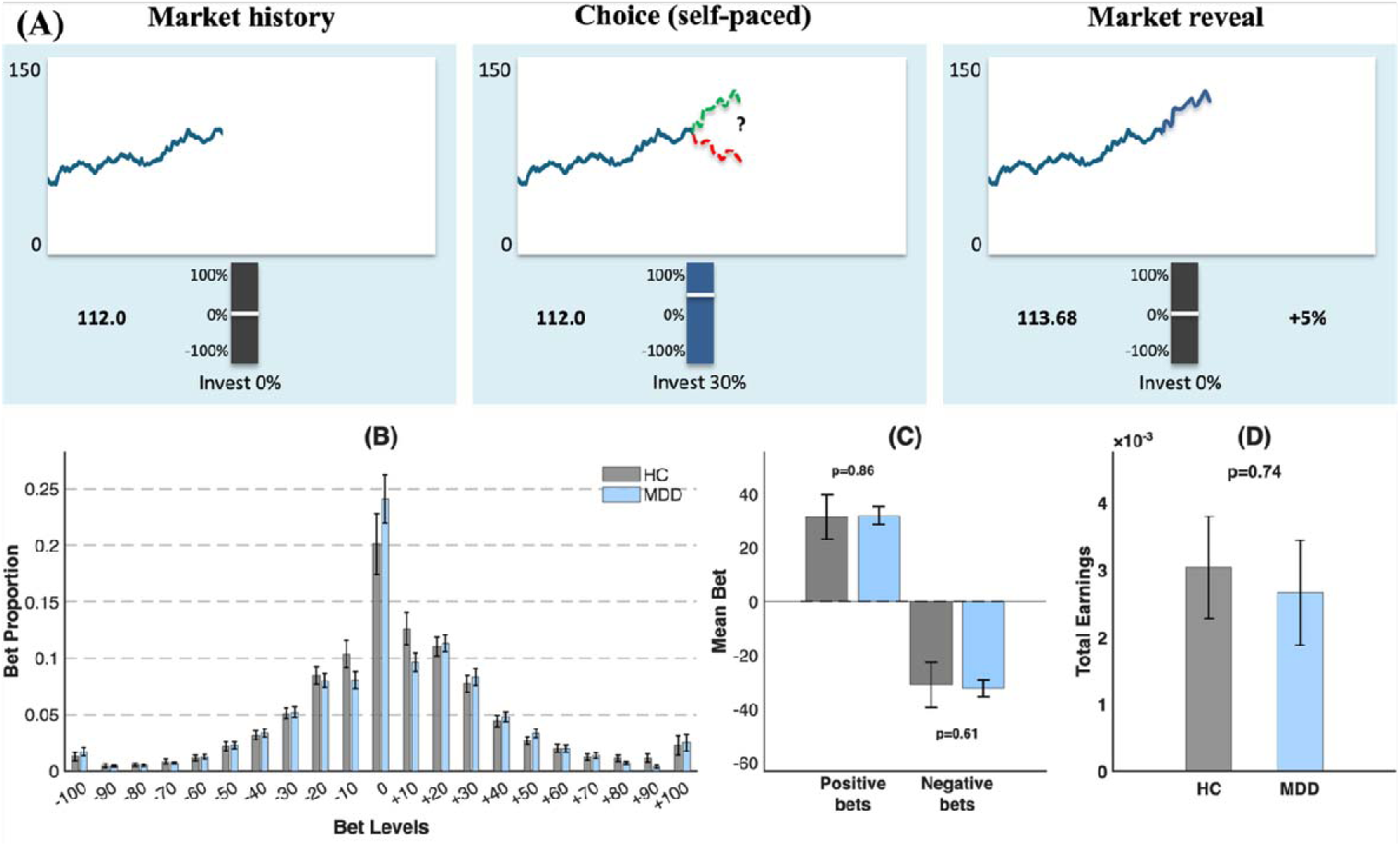
Schematic of the Stock Market Task and summary of betting behavior and earnings. Participants completed 10 markets, each consisting of 20 trials, for 200 trials per session. A trial consisted of three phases (top panel, A). In the Market History phase, participants viewed historical stock price movements. During the Choice phase, they placed a bet on the market’s direction using a slider bar, choosing to invest (positive bet) or short sell (negative bet) a fraction of their portfolio. Green and red dotted lines show possible positive and negative market returns (these lines were not shown to the participant in the task). In the Market Reveal phase, the subsequent market price was displayed, and the participant’s portfolio was updated based on the bet and the fractional return. Because participants could either invest or short, this design separated contextual valence, defined by whether the immediately preceding market return was positive or negative, from instrumental outcome, allowing participants to gain from both market increases and decreases and experience fictive errors across all outcome types. The bottom panel shows (B) Distribution of bet values (proportion of trials) for healthy controls (HC, gray) and participants with major depressive disorder (MDD, blue). Participants exhibited an overall positivity bias, making more positive bets than negative or zero bets. (C) Mean bet amount by bet type by group. Both groups exhibited similar magnitudes for negative and positive bets. (D) Total task earnings did not differ significantly between groups (*p* = 0.74). Error bars represent SEM.

## Results

### Demographic and Clinical Characteristics

Demographic and clinical data are reported in Table 1. As age and race differed significantly between MDD and healthy controls (HC), we conducted a matched-group analysis (see Materials and Methods) and included these variables as covariates in our regression models.

**Table 1.** Demographic and clinical characteristics of the total cohort and fMRI subsample. Data are presented as mean (SD) for continuous variables and n (%) for categorical variables. Statistical comparisons were performed using independent-samples *t*-tests for continuous variables and Pearson’s chi^2 tests (or Fisher’s exact tests for cells with expected counts < 5) for categorical variables. Diagnosis Effect columns display *p*-values for comparisons between Healthy Controls (HC) and Major Depressive Disorder (MDD) patients. Total cohort vs. fMRI subsample columns display *p*-values for comparisons between the total cohort and the nested fMRI subsample to assess potential selection bias. Significant differences (*p* < 0.05) are indicated in bold. Missing data were minimal (<5%) for most variables: HC (n=3 missing for Age, Sex, Education, Race); MDD (n=1 missing for Age, Sex; n=3 missing for Education, Race, SHAPS, QIDS, STICSA). Age of Onset was unavailable for 14 MDD participants. *Abbreviations:* SHAPS, Snaith-Hamilton Pleasure Scale; QIDS-SR, Quick Inventory of Depressive Symptomatology–Self Report; STICSA, State-Trait Inventory for Cognitive and Somatic Anxiety.

|  | Total cohort |  | fMRI subsample |  | Diagnosis Effect (*p*) |  | Total cohort vs. fMRI subsample (*p*) |  |
| --- | --- | --- | --- | --- | --- | --- | --- | --- |
|  | HC | MDD | HC | MDD | Total | fMRI | HC | MDD |
| **Sample size (N)** | 72 | 106 | 26 | 27 | - | - | - | - |
| **Age: Mean (SD)** | 30<br>(10.8) | 36.7<br>(11.8) | 31.2<br>(13.3) | 37.2<br>(12.2) | **< 0.001** | 0.093 | 0.514 | 0.807 |
| **Sex: Female n (%)** | 38<br>(54.3%) | 64<br>(60.4%) | 16<br>(61.5%) | 14<br>(51.9%) | 0.394 | 0.664 | 0.382 | 0.412 |
| **Education** |  |  |  |  | 0.538 | 0.686 | 0.812 | **0.033** |
| Higher education | 24<br>(34.8%) | 30<br>(29.1%) | 10<br>(38.5%) | 12<br>(48%) |  |  |  |  |
| Some college or below | 45<br>(65.2%) | 73<br>(70.9%) | 16<br>(61.5%) | 13<br>(52%) |  |  |  |  |
| **Race** |  |  |  |  | **< 0.001** | **0.01** | 0.20 | 0.82 |
| White/Caucasian | 25<br>(34.7%) | 63<br>(59.4%) | 10<br>(38.5%) | 17<br>(63%) |  |  |  |  |
| Black/African American | 13<br>(18.1%) | 13<br>(12.3%) | 4<br>(15.4%) | 2<br>(7.4%) |  |  |  |  |
| Asian | 28<br>(38.9%) | 13<br>(12.3%) | 12<br>(46.2%) | 4<br>(14.8%) |  |  |  |  |
| Other/Unknown | 6<br>(8.3%) | 17<br>(16%) | 0 (0%) | 4<br>(14.8%) |  |  |  |  |
| **Depression Characteristics** | - |  | - |  |  |  |  |  |
| Medicated (n %) | - | 20<br>(19%) | - | 14<br>(51.9%) | - | - | - | **< 0.001** |
| SHAPS: Mean (SD) | - | 32.2<br>(9.6) | - | 33.5<br>(10.4) | - | - | - | 0.455 |
| QIDS-SR: Mean (SD) | - | 13.6<br>(4.3) | - | 13.3<br>(5.5) | - | - | - | 0.791 |
| STICSA: Mean (SD) | - | 42.4<br>(14.4) | - | 46<br>(13.1) | - | - | - | 0.081 |
| Age of Onset: Mean (SD) | - | 21.1<br>(15.9) | - | 16.1 (9) | - | - | - | **0.04** |
| Psych. Hospitalizations: Mean (SD) | - | 0.9 (5) | - | 2 (9.6) | - | - | - | 0.433 |
| **Comorbid Diagnoses (MDD only)** | - |  | - |  | - | - | - | - |
| GAD | - | 35<br>(34%) | - | 9<br>(33.3%) | - | - | - | ~1 |
| PTSD | - | 8<br>(7.8%) | - | 3<br>(11.1%) | - | - | - | 0.429 |
| Social Anxiety | - | 29<br>(28.2%) | - | 7<br>(25.9%) | - | - | - | 0.959 |
| OCD | - | 3<br>(2.9%) | - | 0 (0%) | - | - | - | 0.565 |

### General Betting Behavior

Across the full cohort, participants showed an overall positivity bias (Figure 1B), placing positive bets more often than negative bets (paired t(177) = 8.81, *p* < .0001). However, baseline betting behavior did not differ between groups: HC and MDD participants showed similar frequencies and magnitudes of positive and negative bets (Figure 1C), overall mean bet size, and task earnings (Figure 1D; all *p* > .25). Detailed descriptive statistics are reported in the Supplement.

### Group Differences in Response to Market Variables

We focus on group differences in how past market experiences influence investment decisions in individuals with MDD versus HC and report the interaction of each task variable with Group (HC, MDD). Complete results from the full regression model are reported in Supplementary Table S1.

### Depression Alters the Influence of Market Return (r) on Bets

Group differences were evident in responses to market fluctuations: specifically, individuals with MDD increased their subsequent bets more than controls did following positive market returns (HC slope = 0.047, *p* < .001; MDD slope = 0.102, *p* < .001; *r^+^* × Group *p* = .038). Conversely, they did not decrease their subsequent bets as much as controls following negative market returns (HC slope = −0.851, *p* < .001; MDD slope = -.738, *p* < .001; *r^-^*× Group *p* = .032), indicating an attenuated behavioral response to market downturns. Crucially, these estimates reflect sensitivity to non-instrumental market trends independent of instrumental (bet-dependent) outcomes: relative to controls, MDD participants were hypersensitive to passive upward market trends yet hyposensitive to passive downward market trends.

### Group Differences in the Influence of Outcome (br) on Bets

Significant group differences were observed in response to outcomes (*br*), specifically under negative contextual valence (Figure 2A). Because market return (*r*) was included as a covariate in the behavioral model, the outcome term captured variance attributable to instrumental fictive error, independent of passive market return. Following invest-losses (*b^+^r^-^*), the MDD group exhibited a significant reduction in bets (MDD slope = −0.577, *p* < .001), whereas HC showed no significant change in their subsequent betting behavior (HC slope = 0.056, *p* = .413; group interaction: β = −0.63, *p* < .001). A similar divergence was observed for short-gains (*b^-^r^-^*). While subsequent bets were strongly and positively associated with short-gains in the HC group (slope = 1.004, *p* < .001), this effect was absent in the MDD group (slope = 0.144, *p* = 0.344; group interaction: β = −0.86, *p* < .001). Since these effects capture the influence of instrumental (bet-dependent) outcomes, the observed divergence indicates a specific breakdown in action-value updating within the MDD group. Together, these results indicate that deficits in counterfactual learning in MDD are uniquely tied to negative contextual valence, characterized by over-updating of bets following losses, reduced adaptation to successful shorting opportunities, and a diminished sensitivity to the passive downward market trend. Under positive contextual valence (Figure 2B), for invest-gains (*b^+^r^+^*), both groups showed significant reductions in subsequent bets (HC slope = −0.48, *p* < .001; MDD slope = −0.359, *p* < .001), with no significant difference observed between groups (interaction *p* = .204). Finally, for short-loss outcomes (*b^-^r^+^*), both groups significantly increased their subsequent bets (HC slope = 0.75, *p* < .001; MDD slope = 0.88, *p* < .001; interaction *p* = .308). Thus, sensitivity to instrumental fictive-error signals appeared preserved in MDD, despite participants being more responsive to the passive upward market trend.

**Figure 2.**
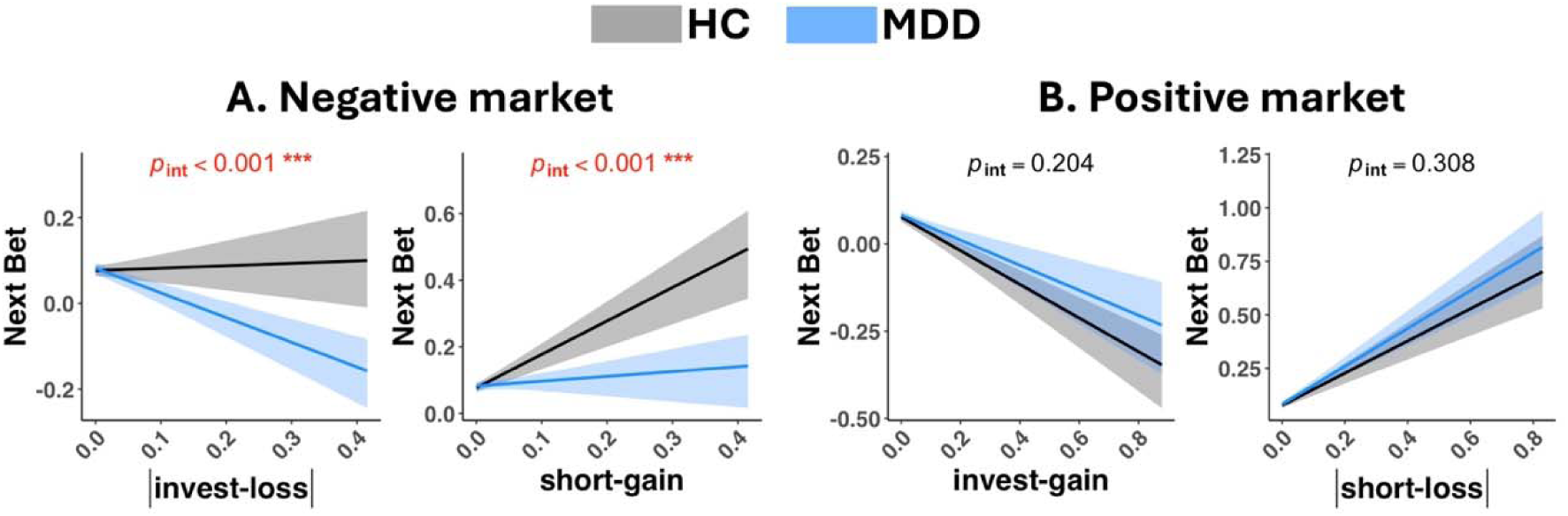
Depression is associated with altered processing of outcomes following negative market returns. Behavioral analysis showing group differences in how participants’ bets changed as a function of outcome from the previous trial. (A) Outcomes following negative market returns: |invest-loss| (b⁺r⁻) and short-gain (b⁻r⁻). (B) Outcomes following positive market returns: invest-gain (b⁺r⁺) and |short-loss| (b⁻r⁺). Healthy control (HC) data are shown in grey, while major depressive disorder (MDD) is in blue. Shaded regions denote 95% confidence intervals. Interaction p-values are shown above each panel. Group differences in betting behavior were context-dependent, emerging only for outcomes following negative market returns. Specifically, MDD patients showed significantly altered sensitivity to invest-loss and short-gain magnitudes compared to HCs, while no significant group differences were observed for outcomes following positive market returns.

Supplementary analyses examining the consistency of these results across behavioral-only and fMRI samples confirmed that the observed clinical group differences were robust across acquisition contexts, as the three-way interactions between diagnosis and market outcomes did not significantly differ between the fMRI and behavioral-only subsamples (all *p* > .055; Table S2). Additionally, the divergence in response to outcomes following negative market returns was robust when comparing only the unmedicated MDD subsample to HCs: invest-loss x Group (β = - 0.727, *p* < 0.001) and short-gain x Group (β = −0.885, *p* < 0.001), and these effects also did not differ significantly by medication status within the MDD group (invest-loss, p = .950; short-gain, p = .177), nor were medication-related differences observed for either positive-market outcome effect (see Supplementary Analyses). However, a significant interaction with diagnosis emerged for short-loss outcomes when comparing the unmedicated MDD subsample to HCs (β = 0.356, *p* = 0.003; Table S2), suggesting that the short-loss effect was specific to the unmedicated subgroup comparison.

### Divergent Neural Processing of Fictive Errors in Depression and Its Clinical Associations

Fictive error signals evoked distinct neural activation patterns across the brain. Invest-gain errors significantly engaged a network including the ACC, bilateral aIns, and VS, whereas invest-loss errors primarily recruited the dACC, aIns, and mid-frontal regions (see Table S4 for peak coordinates). Short-gain outcomes significantly activated the ACC, aIns, and mid-frontal regions, with subthreshold activity also observed in the VS (Table S4). No suprathreshold clusters were observed for short-loss outcomes.

Group-specific activation maps for fictive errors following negative market returns are shown in Figures 3A-B. No group differences in activity survived whole-brain cluster-level FWE correction. ROI analyses revealed a dissociation in the processing of fictive errors following a negative market (Figures 3C-D). For short-gain errors, participants with MDD showed significantly greater activity than the HC group in the dACC (*t* = −2.68; *p*_FDR_ = 0.030) and rACC (*t* = −2.57; *p*_FDR_ = 0.030). The vmPFC showed a similar effect that did not survive FDR correction (*t* = −2.20; *p*_FDR_ = 0.055). While short-gain fictive errors evoked subthreshold VS activation, no significant group differences emerged at the whole brain or ROI analysis levels (NAcc and vCaud *p*_FDR_ > 0.63). For invest-loss errors, there was a trend of increased dACC activation in MDD that failed to survive correction (*t* = −1.97; *p*_uncor_= 0.048).

**Figure 3.**
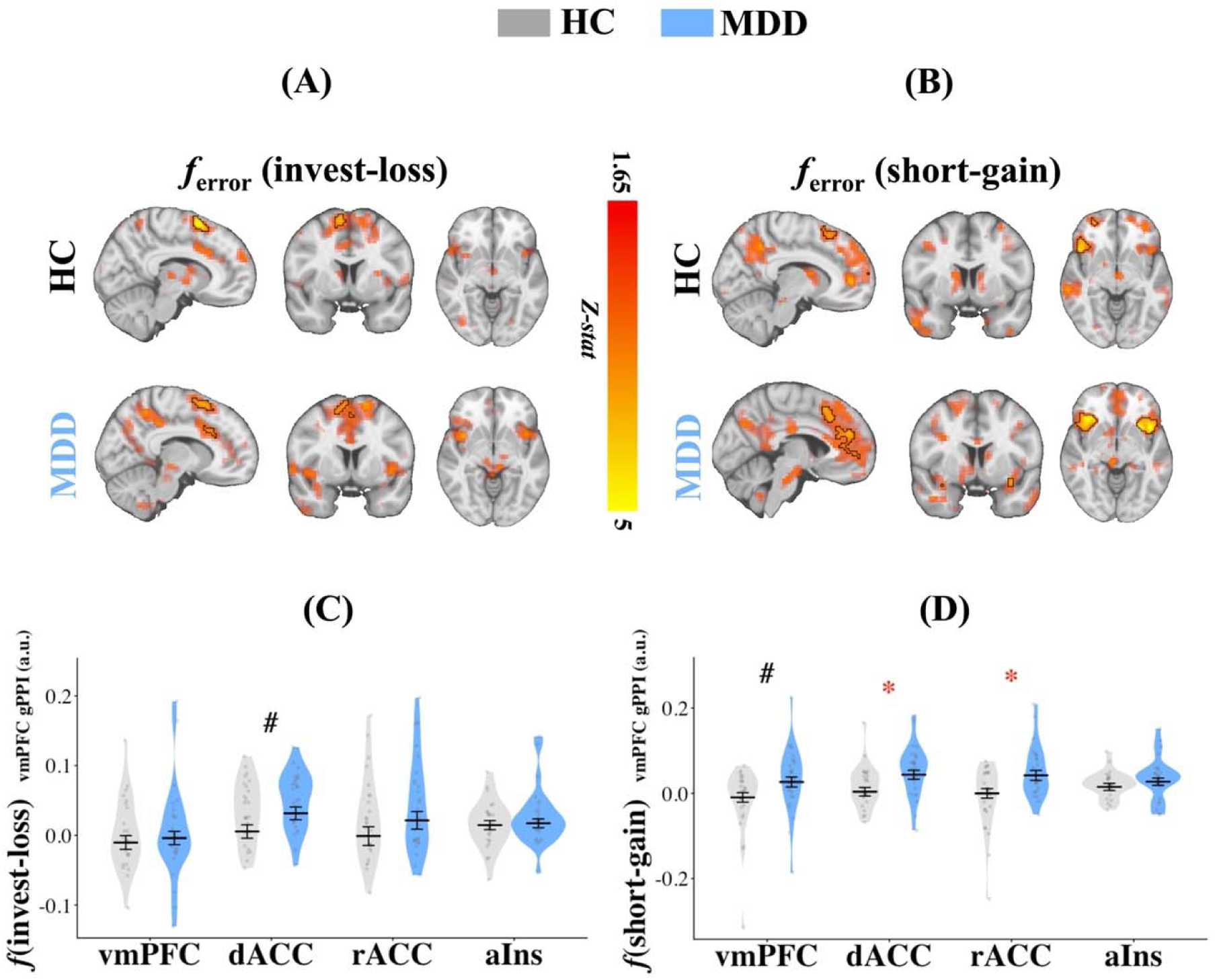
Depression is associated with altered neural processing of fictive errors following negative market returns. **(A–B) Whole-brain activation maps** showing fictive error-related activity (calculated as |r_t_| - b_t_r_t_) for **(A) invest-loss** (b^+^r^-^) and **(B) short-gain** (b^-^r^-^) fictive errors. Separate rows show HC and MDD participants. Opaque regions outlined in black indicate suprathreshold clusters surviving cluster correction at α < .05, using a cluster-forming threshold of p < .001. Faint overlays show subthreshold positive Z-statistics above Z = 1.65 (∼p < .05) for visualization only. Warm colors indicate positive Z-statistics on a common display scale. **(C–D) Region-of-interest (ROI) analysis** quantifying group differences for **(C) Invest-loss** and **(D) Short-gain** fictive error-related changes in BOLD signal. Dots represent covariate-adjusted % change in BOLD signal (controlling for age, sex) derived from a robust linear regression model (weighted by trial count); violin plots show density; error bars indicate mean +/- SEM. For short-gain errors, participants with MDD showed significantly greater activity than HCs in dACC and rACC; vmPFC showed a similar effect that was not significant after FDR correction. For invest-loss errors, a group difference (MDD>HC) was observed in the dACC (*p*_uncor_= 0.048), though it did not survive FDR correction. No significant group differences were observed in the vCaud or NAcc for either error type. Sensitivity analyses confirmed results were unchanged by the exclusion of extreme outliers (defined as values exceeding 3.5 times the IQR; max 2 exclusions per ROI) and remained unmoderated by MDD medication status (all ps > .05). Significance annotations: Asterisks denote False Discovery Rate (FDR) corrected group differences across all six tested ROIs (* *p* < 0.05, ** *p* < 0.01). A black pound sign (#) indicates an uncorrected *p* < 0.05 that did not survive the FDR correction. Abbreviations: vmPFC = ventromedial Prefrontal Cortex; dACC = dorsal Anterior Cingulate Cortex; rACC = rostral Anterior Cingulate Cortex; aIns = anterior Insula; HC = healthy controls; MDD = Major Depressive Disorder.

These group differences in fictive error-related activity following negative market returns were linked to clinical symptoms within the MDD group. For invest-loss (*b^+^r^-^*) fictive error, increased activity in the vmPFC was significantly associated with greater depression severity (β = 0.036; *p*_FDR_ = 0.034), while a similar association in the rACC did not survive correction (β = 0.032; *p*_uncor_= 0.024, *p*_FDR_ = 0.096). For short-gain (*b^-^r^-^*) fictive errors, brain activity was specifically linked to anhedonia-related symptoms rather than general depression. Increased vmPFC activity during short-gain errors was significantly associated with greater anhedonia (SHAPS; β = 0.036; *p*_FDR_ = 0.048) and negatively associated with anticipatory pleasure (TEPS-A; β = −0.032; *p*_FDR_ = 0.048). Additional brain-symptom associations are reported in Table S5. In the positive market context, no associations between symptoms and brain activity survived correction for multiple comparisons (all *p*_FDR_ > 0.07).

### Blunted vmPFC-ACC Connectivity and its Link to dACC Activation Following Negative Market Returns

Task-based functional connectivity (gPPI) analyses, using the vmPFC as a seed (Figure 5A), revealed group differences under negative contextual valence (i.e., following market declines), specifically for short-gain (*b^-^r^-^*) fictive errors (Figure 5B): the MDD group exhibited significantly reduced vmPFC connectivity with both the dACC (*t* = 2.36, *p*_FDR_ = .046) and rACC (*t* = 2.59, *p*_FDR_ = .046) relative to HCs. No significant group differences were observed for invest-loss (*b^+^r^-^*) fictive errors (all *p*_uncor_ > .10; Figure 5C). There were also no significant group differences in fictive-error-modulated connectivity under positive contextual valence (all *p*_uncor_ > .07). Furthermore, connectivity patterns within the MDD group did not differ significantly based on medication status (all ps > .05).

**Figure 4.**
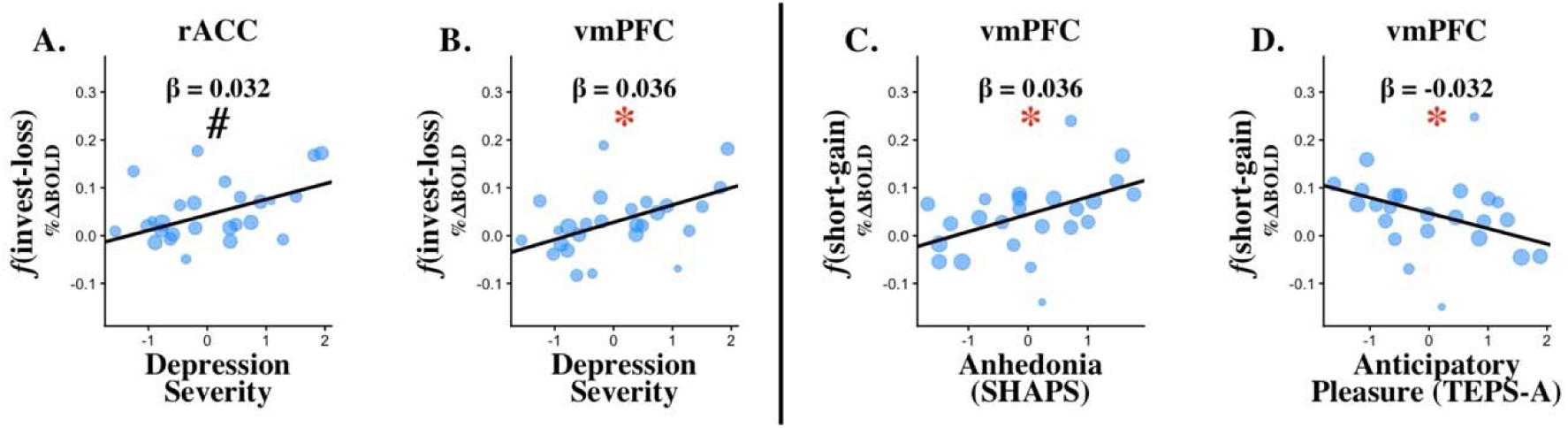
Associations between fictive error-related activity and clinical symptoms following negative market returns. Scatterplots showing the strongest associations (highest effect size) between neural fictive error signals and clinical symptom dimensions, restricted to ROIs that exhibited significant group differences (MDD vs. HC) in the primary analysis. Plots are organized by type of fictive error: (A, B) display responses to invest-loss fictive errors, while (C, D) display responses to short-gain fictive errors. Covariate-adjusted % change in BOLD signal is plotted on the y-axis, against z-scored clinical symptom scores on the x-axis. The solid trend lines represent the fit derived from the robust multiple regression models. Depression severity represents average standardized scores across multiple scales; refer to the “Clinical symptom scores” section for the specific measures used. Standardized beta and *p*-values are annotated on each scatterplot. To control for multiple comparisons, *p*-values were adjusted using the Benjamini-Hochberg procedure (FDR) across the four primary clinical measures (depression severity, SHAPS, TEPS-A, TEPS-C) evaluated within each region of interest and contrast. FDR-corrected *p* < 0.05 is indicated with *, and uncorrected *p* < 0.05 is indicated with #. (A) rACC activity during invest-loss was positively associated with depression severity (uncorrected). (B) vmPFC activity during invest-loss was positively associated with depression severity (FDR-corrected). (C) vmPFC activity during short-gain was positively associated with anhedonia (SHAPS) (FDR-corrected). (D) vmPFC activity during short-gain was negatively associated with anticipatory pleasure (TEPS-A) (FDR-corrected), such that greater activity was related to lower anticipatory pleasure. Models were fit using robust regression, with scatterplot dot size proportional to the observation’s assigned weight (based on behavioral entropy and adjusted trial counts). All models control for age, sex, and medication status. Abbreviations: vmPFC = ventromedial Prefrontal Cortex; dACC = dorsal Anterior Cingulate Cortex; rACC = rostral Anterior Cingulate Cortex; MDD = Major Depressive Disorder; SHAPS = Snaith-Hamilton Pleasure Scale; TEPS-A = Temporal Experience of Pleasure Scale (Anticipatory).

**Figure 5.**
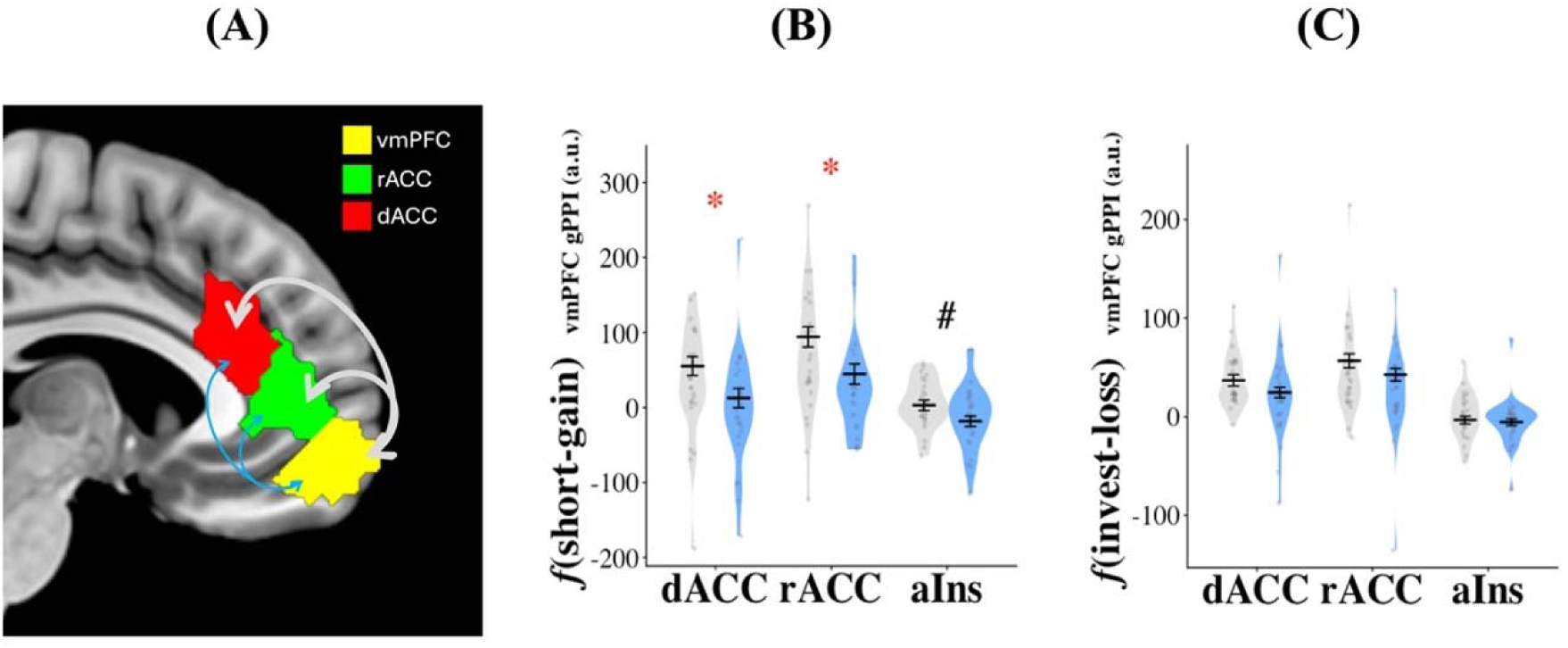
Blunted functional connectivity between vmPFC and the cingulate cortex associated with fictive errors under negative contextual valence in MDD. **A.** The schematic illustrates the task-dependent functional connectivity (gPPI) model, utilizing the vmPFC as the seed region to assess coupling with target ROIs (dACC and rACC shown for simplicity). Line color denotes group (Grey: HCs [HC]; Blue: Major Depressive Disorder [MDD]), and linewidth represents the relative strength of functional connectivity (parameter estimates). **B.** For short-gain outcomes, the MDD group exhibited significantly blunted vmPFC coupling with both the dACC and rACC associated with fictive-error magnitude relative to HCs. A similar reduction in coupling with the anterior insula was observed but did not survive multiple comparison correction. These results were not moderated by medication status. **C.** For invest-loss outcomes, no significant group differences in vmPFC coupling were observed. No significant group differences in connectivity were found across any evaluated ROIs during positive contextual valence. Violin plots illustrate the distribution of connectivity estimates; individual data points represent covariate-adjusted gPPI contrast estimates (a.u.). Thick horizontal lines indicate estimated marginal means; error bars represent +/- 1 SEM. Extreme outliers were excluded using a robust 3.5 IQR threshold (maximum one subject excluded for displayed ROIs). Statistical significance was determined via Type III ANOVA. False discovery rate (FDR) correction was applied across all five evaluated ROI targets. FDR-corrected p < 0.05 is indicated with *, in red, while uncorrected *p* < .05 is indicated using #. Abbreviations: vmPFC = ventromedial Prefrontal Cortex; dACC = dorsal Anterior Cingulate Cortex; rACC = rostral Anterior Cingulate Cortex; aIns = anterior Insula; SEM = Standard Error of the Mean.

This blunted functional coupling during short-gain (*b^-^r^-^*) fictive errors was linked to the hyperactivation observed in the MDD group. Specifically, we found a negative association where lower vmPFC–dACC connectivity predicted higher dACC activity (β = −0.63, *p*_FDR_ = .001). Additionally, during invest-loss (b^+^r^-^) errors, we observed a significant positive association between vmPFC connectivity and anterior insula activity (β = 0.67, *p*_FDR_ = .040). No significant relationships were found between connectivity and activity in the rACC during short-gain errors, nor in either ACC region during invest-loss errors (all *p*_uncor_ > .26). Finally, exploratory analyses revealed no significant associations between task-modulated vmPFC connectivity and clinical symptoms after correcting for multiple comparisons.

## Discussion

Using a novel sequential decision-making task to probe counterfactual processing across positive and negative contexts, we demonstrate that fictive error learning in MDD is not globally impaired but selectively disrupted under negative contextual valence. Specifically, MDD patients were hypersensitive to outcomes that confirmed the negative contextual valence (invest-losses) yet hyposensitive to outcomes that contradicted it (short-gains). This divergent pattern of action-outcome processing was also reflected in diminished sensitivity to passive negative market trends. Neurally, this context-dependent impairment was reflected in altered anterior cingulate and vmPFC activity, as well as reduced vmPFC-cingulate functional connectivity. Together, these results align with a framework of depression in which the negative decision context functions as an overly strong prior, reinforcing outcomes that confirm negative expectations while impairing the integration of successful counterfactual gains into future behavior.

### Negative Contextual Valence Biases Counterfactual Evaluation

The stock market task represents a form of non-stationary reinforcement learning characterized by continuous volatility. This paradigm is akin to probabilistic reversal learning, where participants must learn to switch strategies in response to feedback, albeit in a significantly more volatile environment with no prolonged stable phases. Despite this fluctuating environment, participants successfully tracked the global task dynamics, as the average bet reflected the market’s average return.

Crucially, our results do not support a generalized account of heightened loss learning in depression (15), which would predict a broader increase in behavioral shifting following losses. Instead, patients’ behavioral alterations were dependent on contextual valence, corroborating the notion that reinforcement learning deficits in depression are specifically pronounced in low-value or resource-scarce contexts (37). We suggest this pattern aligns with a decision-theoretic account where overly strong negative priors (i.e., negativity bias) rigidly constrain the updating of action-value beliefs (45). Although patients were less sensitive to the overall passive negative market trend, they showed greater sensitivity to invest-loss outcomes contingent on their bets. Specifically, when outcomes were congruent with the negative environment (invest-losses), MDD participants reduced subsequent bets more strongly than controls, indicating heightened sensitivity to losses under negative contextual valence. This over-adjustment suggests that the “double hit” of sustaining a financial loss within a market downturn reinforced the negative prior, confirming the expectation of loss. Notably, experiencing a loss when the market was going up did not elicit such behavioral alterations in MDD subjects, indicating that the loss oversensitivity here was context dependent.

In contrast, when outcomes were incongruent with negative contextual valence, and subjects made financial gains (short-gains), patients displayed reduced sensitivity to these instrumental outcome signals and failed to capitalize on the opportunity to optimize future choices. Again, the MDD group did not show behavioral deficits in processing gain outcomes during trials with positive market returns, and their deficits were limited to the negative decision context. This behavioral pattern is consistent with mood-congruent pruning of actions (46), where dominant negative priors inhibit the consideration of rewards. Our findings extend previous work by showing that reduced sensitivity to rewards may depend on the decision context and that an aversive context may lead to the elimination of potentially profitable action paths (45). Together with our findings related to heightened sensitivity to loss outcomes during the negative market return trials, these results reflect a context-dependent bias rather than a global pruning deficit (47).

Although adapting behavior based on counterfactuals recruits evaluative processes akin to model-based thinking – requiring internal representation of unrealized outcomes and enabling prospective regret-averse behavior informed by current fictive error signals – our results suggest that depression is not characterized by a generic impairment in such reasoning. Notably, depressed participants successfully utilized fictive error information to guide decisions in positive market contexts, performing similarly to healthy controls. Instead, our results align with evidence that a negatively valenced environment hampers model-based thinking in depression (48). Specifically, while patients showed blunted updating in response to positive counterfactuals (short-gains), they reinforced negative response tendencies following losses, effectively over-learning from outcomes that confirmed their bias (49).

### Altered Frontocingulate Processing Under Negative Contextual Valence

The behavioral pattern observed in MDD under negative contextual valence was mirrored by distinct neural patterns. The strongest group differences emerged during short-gain fictive errors, for which MDD participants exhibited significantly greater activity than controls in the dACC and rACC. Although group differences in vmPFC activity did not survive correction, vmPFC activity correlated with depression and anhedonia symptoms during trials following negative market returns. This pattern is broadly consistent with prior evidence of atypical activation in these regions during reward assessment in MDD (16, 44, 50, 51). These regions have been shown to have overlapping but distinct roles in cognition: while the vmPFC serves as an integration hub for computing relative value (52), the dACC utilizes these inputs to decide on actions and assess their outcomes (53–55). The rACC, like the vmPFC, is a part of the default-mode network (56) and is implicated in affective rumination (51, 57).

While reports of frontocingulate activation in MDD include both under- and over-recruitment, the heightened dACC and rACC responses observed here may reflect inefficient engagement of frontocingulate systems during short-gain fictive errors (51, 58). Specifically, this pattern parallels a profile of failed deactivation in default-mode regions (rACC/vmPFC) occurring alongside hyperactivation in cognitive control regions (dACC), suggesting that depressed individuals inefficiently recruit cognitive control networks in an effort to counteract the interference generated by negative information (56, 59). However, the clinical significance of these neural signals dissociated by condition. For invest-loss fictive errors, vmPFC activation increased with depression severity. This suggests that, in more severely depressed individuals, outcomes that confirm a negative context engage valuation-related processing more strongly. Conversely, neural responses to short-gains specifically tracked hedonic deficits: heightened vmPFC activity scaled with anhedonia (SHAPS) and varied inversely with anticipatory pleasure (TEPS-A), while similar associations in the rACC did not survive FDR correction. Together, these findings map a dual-process failure in which the depressive brain is hypersensitive to context-congruent losses yet hyposensitive to context-incongruent gains.

Task-based functional connectivity analysis revealed that during context-incongruent gains (short-gains) in negative markets, depressed participants exhibited significantly blunted coupling between the vmPFC and ACC subregions (rACC and dACC). The ACC receives projections from the vmPFC (60), providing a pathway through which value-related and action-relevant information may influence behavioral adjustment. Emerging theories propose that the vmPFC contributes to representing value based on internally simulated outcomes (61, 62), as well as signaling the extent to which outcomes are contingent on one’s own actions (63, 64). In the stock market task, tracking fictive error likely depends on this type of computation: evaluating actual outcomes while mentally simulating alternate outcomes that could have happened. Attenuated functional connectivity between these regions in MDD (65, 66) scales with depression severity (67) and may contribute to difficulty overcoming negative processing biases. In our study, this decoupling may reflect reduced transmission of vmPFC signals related to value, agency, or perceived control (68) when outcomes contradict negative contextual valence, consistent with evidence that fronto-cingulate coupling helps regulate the subjective impact of regret (36). During short-gain fictive errors, lower vmPFC–dACC connectivity also predicted higher dACC activity. This negative association suggests that greater dACC recruitment occurs when coordination with vmPFC valuation-related signals is reduced. A similar dissociation between fictive error-related neural signaling and its behavioral impact has been noted in chronic smokers, where fictive error signals were neurally encoded but failed to guide subsequent choice, while rACC activity tracked subjective motivational state (i.e., craving) rather than adaptive choice behavior (5). Collectively, these findings converge on the role of cognitive appraisal in regulating fictive error-related affect and adaptation (11, 36), and may help explain prior reports of a disconnect between subjective regret and behavioral adaptation in depression (38, 40).

### Limitations

Several limitations should be considered. First, the MDD group differed from controls in age and racial composition. However, our core findings remained robust after correcting for these imbalances using a matched iterative sampling approach, suggesting demographic variables did not drive the results. Second, while behavioral shifts were interpreted as regret-driven, the task did not collect trial-by-trial subjective ratings of regret. Consequently, without direct measures of felt emotion, we cannot definitively map the observed neural signals to conscious emotional states, which is needed to explicitly explore any disconnect in affect and behavioral adaptation. Finally, a significant portion of the fMRI population was medicated. However, we observed no differences in neural or behavioral markers between medicated and unmedicated subgroups, suggesting the circuit disruptions may represent a stable trait, consistent with evidence that frontocingulate dysfunction persists even in remission (69). Notably, while pharmacological interventions typically improve reinforcement learning in a valence-independent manner (70), the core deficits under negative contextual valence identified here remained unmoderated. Medication did, however, appear to be associated with a reduced short-loss abnormality, as this effect was evident only in the unmedicated MDD subgroup relative to HCs. This pattern is consistent with the possibility that treatment may facilitate specific adaptive shifts without fully remediating the primary hypersensitivity to negative environments. However, given our cross-sectional design and small medicated sample (N = 20), these observations remain tentative and require further validation.

### Conclusion

We demonstrate that impairments in counterfactual learning and behavioral updating in MDD are not global but emerge specifically through an interaction with contextual valence. Behaviorally, patients exhibited a dual failure: hypersensitivity to outcomes that confirmed negative contextual valence (congruent losses) and hyposensitivity to those that contradicted it (incongruent gains). Neurally, the clearest abnormalities were observed during context-incongruent gains, which were associated with heightened dACC and rACC responses and reduced vmPFC-cingulate functional connectivity. During context-congruent losses, vmPFC activity tracked depression severity within the MDD group. Therapeutic strategies should therefore take into consideration the broader context of learning, as divergent approaches may be needed to restore adaptive behavior depending on the environment.

## Materials and Methods

### Participants

Participants included individuals with major depressive disorder (MDD) and healthy controls (HC), 18-75 years of age. Behavioral data were collected from 178 participants, recruited at either The University of Dallas, Texas (between 2015 and 2018) or Icahn School of Medicine at Mount Sinai (between 2021 and 2024), all of whom completed an identical stock market investment task. The Institutional review boards at both institutions approved the study, and written informed consent was obtained from all participants prior to any study procedure. Individuals in the MDD group met Diagnostic and Statistical Manual of Mental Disorders (DSM-5) (71) criteria for major depressive disorder (MDD), as determined by the Structured Clinical Interview for DSM-5 Disorders (SCID). HC participants had no history of any psychiatric disorder, as confirmed by the SCID. Participants were not required to discontinue any current medications (including antidepressants) or therapies for this study. Participants were excluded if they had a history of any psychotic, neurodevelopmental, or neurocognitive disorders. Additional exclusion criteria included an active substance use disorder within the past year or a positive urine toxicology screen at screening. All participants provided informed consent. A subset of participants (n=53) completed the task while undergoing functional magnetic resonance imaging (fMRI) at the Icahn School of Medicine at Mount Sinai. For these fMRI participants, contraindications to MRI were imposed as additional exclusion criteria. Key demographic and clinical characteristics for both the full behavioral sample and the fMRI subset are presented in Table 1.

### Stock market task

This novel task was developed based on a previous sequential investment task (5, 6, 11, 72, 73). Participants were allocated an initial sum of 100 monetary units (i.e., their portfolio) at the beginning of the experiment, which they could use to place bets (Figure 1) in the stock market. Participants were informed that their final payment would include a bonus that was scaled according to their actual gains or losses in the task. Each task block commenced with a caption titled ‘new market’ followed by a graphic display of past market dynamics. In each trial t, the participant observed the price history of a stock market (including the trial before, p_t-1_) and placed a bet, b_t_. Distinct from previous studies, participants were able to choose to either make positive bets (*inves*t; if they predict the market price will go up) or negative bets (*short*; if they predict the market price will go down). Notably, shorting the market would result in gains from market drops. Thus, people could benefit from either a positive or negative price change. Participants provided their bets using a slider bar and finalized their choice by a button press. Following a 750-ms delay, the new market price was p_t_ was revealed, and portfolio amounts were updated to reflect the recent outcome. The fractional market return r_t_ is defined as r_t_ = (p_t_ – p_t-1_) / p_t-1_. The outcome from a trial was therefore b_t_r_t_. Crucially, this design dissociates the general passive market return from the instrumental outcome: a positive market does not necessarily result in a gain, nor does a falling market imply a loss. This allowed for the independent assessment of behavioral sensitivity to non-instrumental market trend r_t_ versus evaluative feedback – specifically the realized outcome b_t_ r_t_ and the associated fictive error (r_t_ - b_t_ r_t_) resulting from the participant’s instrumental choice.

Participants had unconstrained time to decide on their investments. Thus, natural variability in participants’ response times generated time jittering between trials. Each participant played a total of 10 markets, each market consisting of 20 trials. The stock market prices in the task were chosen from true historical stock market prices. A total of 30 different markets were used across all participants, with the 10 markets for a specific participant randomly sampled from this pool of markets. Prior versions of the task (with healthy participants) that allowed only positive (invest) bets (and therefore a gain in only positive markets) demonstrated that following invest–gain outcomes, higher gains (i.e., lower fictive errors) typically led to lower subsequent investments (5, 6, 11), consistent with regret-averse behavior (1, 6). See Supplementary Figure S1 for hypothesized patterns of bet adjustment based on regret-averse behavior in this task.

Participants with flat or close to flat responses (bets) were excluded based on Shannon entropy (74) (minimum possible entropy = 0 if all investment responses are the same across trials; maximum possible entropy = log2(n), where n = the number of possible responses), calculated for each individual’s bet responses. Eight participants (three MDD, five HC) with entropy scores ≤2 standard deviations below the sample mean were excluded from all analyses.

### Clinical symptom assessment

General depression severity was assessed using the Quick Inventory of Depressive Symptomatology–Self Report (QIDS-SR (75)) and the depression subscale of the Depression Anxiety Stress Scale (DASS-D (76)). Subjects also completed the Snaith-Hamilton Pleasure Scale (SHAPS) and the Temporal Experience of Pleasure Scale (TEPS (77)) as measures of anhedonia. The TEPS was subdivided into anticipatory (TEPS-A) and consummatory (TEPS-C) subscales to capture reward ‘wanting’ and ‘liking’ components (78), respectively. Anxiety was evaluated using a comprehensive composite including the DASS-Anxiety (DASS-A), the State-Trait Inventory of Cognitive and Somatic Anxiety (STICSA(79); cognitive and somatic subscales), the State-Trait Anxiety Inventory (STAI (80); state and trait versions), and the MASQ-Anxious Arousal (MASQ-AA (81)). Certain clinical symptom measures were not collected for a subset of participants in the behavioral sample (see Table 1), reflecting site-specific differences in assessment protocols.

### Image acquisition and preprocessing

All MRI data were acquired with a Siemens 3T MAGNETOM Skyra scanner and a 32-channel head coil. Scans included an anatomical T1-weighted scan and a functional scan of the Market task. The anatomical T1-weighted images were acquired with a magnetization-prepared rapid gradient-echo sequence, repetition time (TR)□=□2400□ms, echo time (TE)□=□2.07, inversion time□=□1000 ms, field of view (FOV)□=□320□×□320, voxel resolution□=□0.8□×□0.8□×□0.8□mm^3^), and the functional scan for Market task performance was collected with a multi-echo (ME) multi-band accelerated echo planar imaging sequence (TR□=□882□ms, TEs□=□11.0, 29.7, 48.4, 67.1, multi-band factor□=□5, FOV□=□560□×□560, voxel resolution□=□3□×□3□×□3□mm^3^, flip angle□=□45).

Functional scans were preprocessed and denoised for motion and physiological noise using multi-echo-independent component analysis (ME-ICA (82)). In this pipeline, AFNI (83) is used, for each echo, to conduct despiking (*3dDespike*), correction for slice timing offsets (*3dTshift*), estimating and applying motion-correction realignment (*3dvolreg*) using the middle echo by aligning it to the minimum-outlier volume. The multiple echoes are then combined using a weighted-average approach, balancing the higher signal from earlier echoes with the greater sensitivity of later echoes. The resulting whole-brain time series is then dimensionally reduced using principal component analysis (PCA), followed by independent component analysis (ICA) to remove non-BOLD components. Components with high echo-time dependence are considered BOLD-like and accepted, while those with low dependence are noise-like and rejected (84). This method facilitates robust denoising for motion, physiological, and scanner artifacts (84, 85). Denoised functional data were co-registered with their respective T1 and normalized to a standard Montreal Neurological Institute (MNI) template using ANTS (86) and spatially smoothed with a 6□mm full-width-at-half-maximum kernel.

### Behavioral modeling

The effect of market variables from previous trials on the participant’s bet decision in the subsequent trial was assessed using linear mixed-effects multiple regression analyses. Mixed-effects models were estimated in R (RStudio (87)) using the ‘lmer’ function. The model specification was as follows:

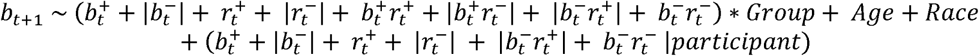

Where Group is a factor with two levels, HC and MDD. Importantly, by including market return (*r*) as a covariate, the outcome (*br*) term uniquely captures the variance attributed to the instrumental fictive error (|*r*| - *br*). The final return of each market was excluded from the regression, as there is no investment decision following the final market segment. Similarly, the first trial was also removed since it had no preceding investment decision. Linear mixed effects were estimated using the Restricted Maximum Likelihood (REML) method.

Model structure was determined through iterative optimization (88). The final random-effects specification included slopes for positive and negative bets, positive and negative market returns, and the two shorting outcome types. To address baseline group differences in Age and Race (see Table 1), we conducted a matched-group analysis using MatchIt (89) to balance the groups based on Age, Race, and Education. Following this matching procedure, to account for unbalanced sample sizes between groups, we calculated confidence intervals and *p*-values for the effects using an iterative resampling method with 15,000 iterations (see Supplementary Materials for more details). Additionally, to rigorously separate core MDD deficits from potential confounds, we conducted additional supplementary sensitivity analyses: (1) generalizability test (Task Variables x Group × Acquisition Context) to determine if key results differed significantly between the behavioral-only and fMRI subsamples; (2) a matched-group analysis restricted to unmedicated MDD to remove pharmacological confounds; and (3) a within-MDD assessment of medication effect.

### General linear modeling (GLM) of fMRI data

We specified two primary first-level GLMs for each subject. In both GLMs, we included the following regressors: (i) new market display, (ii) market history display, (iii) all key presses, (iv) market price reveal of round 1, (v) market price reveal of rounds 2–19, and (vi) market price reveal of round 20. Regressors were constructed by convolving events at the onset of each stimulus or motor response with the double-gamma hemodynamic response function (HRF) implemented within FSL (90). Additional regressors were constructed from the markets and bets. In the first “market” model, the percentage of market return, r⁺ (positive market returns) and |r⁻| (absolute value of negative market returns) parametrically modulated the time of market reveal. In a second “fictive error” model, the market-reveal events were parametrically modulated by the four types of fictive error regressors. The fictive error (*f*) regressor was calculated as |r| - br (see Figure S1), reflecting the magnitude of relative loss or “regret” (6).

### Generalized psychophysiological interaction (gPPI)

Task-based functional connectivity was assessed using generalized psychophysiological interaction (gPPI) (91). To investigate task-dependent functional coupling during counterfactual processing, we selected the vmPFC as the seed ROI given its role as a key node in regret-related valuation and appraisal networks (43, 61), its broader relevance to prefrontal dysfunction in depression (59), and a nominal effect in the primary GLM (see ROI definition in Figure S2). For each participant, the denoised BOLD time series was extracted from the seed ROI and deconvolved using a Wiener filter to estimate underlying neuronal activity (92). Interaction terms were generated by multiplying this estimated neuronal signal with the unconvolved task regressors (onset times and durations, with parametric modulators demeaned). The resulting neural interaction vectors were then convolved with a double-gamma hemodynamic response function (HRF). The first-level gPPI GLM included the convolved task regressors (psychological variables), the seed ROI time series (physiological variable), and the convolved interaction terms. Functional connectivity with the vmPFC seed was subsequently evaluated across all previously defined target ROIs.

### Whole brain group-level analyses

First-level GLM contrast images for each participant were entered into group-level analyses to identify overall (HC+MDD), group-specific (HC only, MDD only), and group-difference (HC vs. MDD) activations. Group-level analyses were performed using FLAME1 in FSL (93), with age and sex included as covariates. Significant activations were determined using cluster-extent thresholding (94), with clusters defined by Z□>□3.09 (uncorrected p□<□.001) and a corrected cluster significance threshold of p□<□.05 (95).

### ROI Selection and Signal Extraction

We defined regions of interest (ROIs) based on their established involvement in reward-related computations—particularly the encoding of experiential and fictive error signals (using this task: (5, 6, 11, 72, 96); also fictive-error/regret meta-analysis (43)) – as well as their involvement in the manifestation of depression (97–101). Regions of interest (ROIs) were defined using standardized, unbiased atlases. The ventral caudate (vCaud) and nucleus accumbens (NAcc) were derived from the Harvard-Oxford subcortical atlas in FSL, with the vCaud ROI further restricted to caudate voxels inferior to z = 7 (MNI coordinates), following prior work (102). Cortical ROIs, including the dorsal and rostral anterior cingulate cortices (dACC, rACC), anterior insula (aIns), and the ventromedial prefrontal/medial orbitofrontal cortex (vmPFC/mOFC), were selected from the Schaefer functional parcellation atlas (103) (see Figure S2).

GLM-derived contrast parameter estimates (COPE) were extracted and converted to percent signal change (PSC; %ΔBOLD) for each ROI to facilitate comparison across subjects. PSC was calculated by normalizing (dividing) the COPE value by the mean of the functional signal and scaling it by the group-level average peak-to-peak height of the effective regressor for the contrast (104).

### Group Differences and Brain–Symptom Associations

Group differences in ROI activity and their associations with clinical symptoms within the MDD group (for ROIs demonstrating significant group differences) were evaluated using robust regression models weighted by condition-specific trial counts to account for measurement reliability. All models controlled for age, sex, and medication status. To reduce dimensionality and capture common variance, symptom domains comprising multiple instruments (depression and anxiety) were created by individually standardizing (z-scoring) constituent measures and then averaging them. For comparisons between groups, univariate outliers were identified and excluded if ROI values exceeded 3.5 times the interquartile range (IQR) within each ROI. False discovery rate (FDR) correction was applied across the six ROIs for tests of group differences in ROI activity and connectivity, and across the four key symptom measures for brain-symptom analyses.

## Data Availability

All data produced in the present study are available upon reasonable request to the authors.

## Acknowledgments

Nil.

