## Supplementary for "Negative Contextual Valence Unmasks Altered Counterfactual Decision-Making in Major Depressive Disorder"

**Supplementary Material**

**Demographics differences**

When comparing the total cohort to the fMRI subsample, the subsample was largely representative of the total group regarding Age, Sex, and Race. However, a significant difference was found in Education level (p = 0.033). Medication Status was substantially higher in the fMRI subsample (51.9% vs. 19% in the total MDD cohort; p < 0.001), with a difference in age of illness onset in the fMRI subsample (16.1 years vs. 21.1 years in the total MDD cohort; p = 0.040). Within the total MDD cohort, the most common comorbidities were Generalized Anxiety Disorder (GAD, 34%) and Social Anxiety (28%).

Consistent with prior work distinguishing fMRI from behavioral-only acquisition settings (1, 2), betting patterns differed significantly between acquisition context. The fMRI group made significantly fewer (22.8% vs. 31.3%; *p* < 0.001) and lower magnitude (*p* = 0.025) negative bets, replacing them with zero bets (37.4% for fMRI vs. 28.3% for behavioral-only; *p* = 0.006), while positive betting behavior remained stable in both frequency (*p* = 0.82) and magnitude (*p* = 0.07). These acquisition-related shifts were not significantly modulated by diagnosis (all interaction *p*s > .05).

**General Betting Behavior and Earnings**

Despite the equal possibility of placing positive and negative bets, participants bet more positively overall (mean bets = 4.91%, *p* < 0.0001, 95% CI [3.54, 6.27]; number of positive bets 45.2% of trials, negative bets 32.3% of trials; paired *t*(177) = 8.81, *p* < 0.0001), demonstrating a positivity bias. Overall betting frequencies remained balanced between diagnostic groups in the total cohort. The frequency of positive bets was similar across groups (HC: 46.3%, MDD: 44.4%, *t*(176) = 0.76, *p* = 0.45), as was the frequency for negative bets (HC: 33.6%, MDD: 31.5%, *t*(176) = 1.03, *p* = 0.30). No significant difference was found for zero bets (HC: 20.1%, MDD: 24.1%, *t*(176) = –1.17, *p* = 0.25).

Linear mixed-effects models with subject as random effect showed that both groups exhibited similar magnitudes for negative bets (MDD: -32.27% vs HC: -30.94%, *p* = 0.61) as well as for positive bets (MDD: 32.00% vs HC: 31.54%, *p* = 0.86), and overall mean bet amount did not differ by group (MDD: 4.74% vs HC: 5.15%, *p* = 0.78). Average bet amount for a specific market was significantly predicted by that market's average return (β = 1.18, *t*(1759) = 5.73, *p* < 0.0001), and this relationship did not differ significantly between groups (interaction *p* = 0.18). Task earnings did not differ between groups (HC: 0.30 ± 0.65, MDD: 0.27 ± 0.80; *t*(176) = 0.33, *p* = 0.74, 95% CI [-0.19, 0.26]). Mean session duration in the stock market task was 13.87 ± 4.47 min and did not differ between groups: t (132) = 0.55, *p* = 0.59.

**1. Model Selection Strategy**

We used an iterative approach to determine the optimal random effects structure. The maximal model including all eight random slopes (the four market variables: ***b^+^***, ***b^-^***, ***r^+^***, ***r^-^***; plus outcomes: ***b^+^r^+^***, ***b^+^r-***, ***b^-^r^+^***, ***b^-^r^-^***) by participant yielded a singular fit, indicating overparameterization. We therefore compared two simplified structures: a model with only the four main effect slopes (Model 1: AIC = 14,173) versus a model with only the four interaction slopes (Model 2: AIC = 15,216). Model 1 showed superior fit, serving as our baseline for further refinement.

In Phase 1, we systematically tested adding each outcome type-related random slope individually to Model 1. Inclusion of the ***b^-^r^+^*** outcome as random slope showed the largest improvement (change in AIC = -399, AIC = 13,774), followed by ***b^-^r^-^*** (change in AIC = -219), ***b^+^r-*** (change in AIC = -202), and ***b^+^r^+^*** (change in AIC = -226). In Phase 2, we tested adding a second outcome-related slope to the best Phase 1 model. Adding ***b^-^r^-^*** (AIC = 13,636, change in AIC = -537 from Model 1) produced clean convergence, while adding ***b^+^r-*** (AIC = 13,604) yielded only marginal additional improvement (32 AIC points) but did not converge well.

We selected the model with main effect slopes plus both short-selling outcome type slope terms (***b^-^r^+^*** and ***b^-^r^-^***) as our final model, given its substantially improved fit over the baseline, clean convergence, and theoretical coherence. The importance of short-selling interaction slopes is consistent with our observation that participants shorted less frequently (32.3% of trials) than they invested in positive positions (45.2% of trials), with short-selling showing greater context-sensitivity and between-person variability in response patterns. Final random effects structure: (***b^+^*** + ***b^-^*** + ***r^+^*** + ***r-*** + ***b^-^r^+^*** + ***b^-^r^-^*** | subject)

**2. Matched-Group Analysis and Iterative Inference**

To account for demographic imbalances in Age and Race (Table 1), we utilized the MatchIt package (3) to implement 1:1 optimal matching on Age, Race, and Education, with an exact match constraint on the dichotomized Race variable (White vs. Non-White). Post-matching diagnostics, using *t*-tests for Age and Chi-square tests for Race, confirmed that groups were balanced and statistically indistinguishable across all matched covariates (p > .05). Final statistical inference was conducted using an iterative resampling approach consisting of 15,000 iterations. To ensure the results were not dependent on specific participant combinations, each iteration randomly assigned either the Healthy Control (HC) or Major Depressive Disorder (MDD) group for 90% sub-sampling, thereby introducing matching variance. Each iteration matched samples consisting of 61 to 64 pairs of participants (i.e., a total N ranging from ~ 122 to ~ 128), depending on the subsampling. Following each matching step, we fitted a linear mixed-effects model using the bobyqa optimizer. Only iterations that achieved both successful matching and model convergence were included in the final analysis (~ 99% of iterations). To validate the diversity of the matched samples, we calculated the Jaccard Distance (Mean Variation) across all successful iterations. Within-strategy variation was 17.6%–18.5% - confirming that the randomized sub-sampling successfully introduced sufficient diversity into the samples. Final p-values and empirical 95% intervals were derived from the resulting distribution of fixed-effect coefficients across successful iterations. For within-group effects, slopes for the reference group (HC) were taken directly from the fixed-effect coefficients, whereas MDD slopes were derived within each iteration by adding the corresponding Group × Predictor interaction term; final group-specific slopes were then averaged across iterations. For all fixed effects, including main effects and Group × Predictor interactions, p-values were calculated as twice the smaller proportion of the empirical coefficient distribution falling above or below zero. For within-group simple slopes, slopes for the reference group (HC) were taken directly from the fixed-effect coefficients, whereas MDD slopes were derived by adding the corresponding Group × Predictor interaction term; the same empirical p-value procedure was then applied to these group-specific slope distributions.

| **Table S1. Linear-mixed-effect results of the primary behavioral model** | | | | |
| --- | --- | --- | --- | --- |
| **Effect** | **Beta** | **CI_lower** | **CI_upper** | **Empirical_p** |
| (Intercept) | 0.077 | 0.064 | 0.089 | < .001*** |
| ***b^+^*** | 0.310 | 0.282 | 0.341 | < .001*** |
| ***b^-^*** | -0.352 | -0.375 | -0.328 | < .001*** |
| ***r^+^*** | 0.047 | 0.025 | 0.081 | < .001*** |
| ***r-*** | -0.851 | -0.945 | -0.779 | < .001*** |
| ***b^+^r^+^*** | -0.482 | -0.623 | -0.384 | < .001*** |
| ***b^+^r-*** | 0.056 | -0.183 | 0.326 | 0.413 |
| ***b^-^r^+^*** | 0.752 | 0.558 | 0.949 | < .001*** |
| ***b^-^r^-^*** | 1.004 | 0.669 | 1.265 | < .001*** |
| GroupMDD | 0.006 | -0.002 | 0.013 | 0.150 |
| Age | -0.001 | -0.001 | 0.000 | 0.001*** |
| Race | -0.004 | -0.011 | 0.003 | 0.237 |
| **Group interaction effects** | | | | |
| ***b^+^***:GroupMDD | -0.029 | -0.069 | 0.010 | 0.144 |
| ***b^-^***:GroupMDD | 0.053 | 0.014 | 0.093 | **0.007**** |
| ***r^+^***:GroupMDD | 0.055 | 0.004 | 0.103 | **0.038*** |
| ***r-***:GroupMDD | 0.114 | 0.009 | 0.226 | **0.032*** |
| ***b^+^r^+^***:GroupMDD | 0.123 | -0.070 | 0.311 | 0.204 |
| ***b^+^r-***:GroupMDD | -0.633 | -0.926 | -0.345 | **< .001***** |
| ***b^-^r^+^***:GroupMDD | 0.132 | -0.120 | 0.394 | 0.308 |
| ***b^-^r^-^***:GroupMDD | -0.860 | -1.284 | -0.441 | **< .001***** |
| **Note:** Linear mixed-effects model with next bet (bₜ₊₁) as the dependent variable and the following fixed-effect regressors: b⁺ and b⁻, prior investment decisions (positive and negative bet, respectively); r⁺ and r⁻, positive and negative market returns respectively; b⁺r⁺ and \|b⁺r⁻\|: outcome magnitudes for invest-gain and invest-loss, respectively. \|b⁻r⁺\| and b⁻r⁻: outcome magnitude for short-loss and short-gain, respectively. Interaction terms with Group are included to examine group-specific effects. Main effects are conditional effects relative to the reference group (healthy controls, HC). For numeric predictors, estimated slopes reflect effects within the HC group. Interaction terms with Group tested whether the relationship between each predictor and the outcome differed between major depressive disorder (MDD) and HC participants. **Statistical inference:** To ensure robustness of results and demographic control, final statistical inference was conducted using an iterative ensemble matching procedure (15,000 iterations). Each iteration involved optimal 1:1 matching followed by linear mixed-effects model (LMM) fitting. The reported beta coefficients and 95% confidence intervals (CI) represent the mean and empirical distribution of estimates across all iterations. The *p*-value for each effect was derived from the empirical distribution, calculated as twice the minimum proportion of the distribution of coefficients that lay on either side of zero. LMM was fitted using restricted maximum likelihood (REML), with the *random effects structure:* (***b^+^*** + ***b^-^*** + ***r^+^*** + ***r-*** + ***b^-^r^+^*** + ***b^-^r^-^*** \| subject). Significance levels: **p* < 0.05, ***p* < 0.01, ****p* < 0.001. | | | | |

**Supplementary Sensitivity Analyses**

**Supplementary Analysis 1: Generalizability of fMRI vs. Behavioral-only data**

Given that shorting behavior differed significantly between the fMRI and behavioral-only samples (e.g., reduced short-selling frequency in the scanner), we tested whether the diagnostic group differences observed in the full sample were stable across acquisition contexts. We fitted a full-cohort interaction model that included Task Variable x Group x Context interaction term (where Context was a binary factor: fMRI vs. Behavioral-Only). A non-significant 3-way interaction indicates that the difference between MDD and HC participants remains consistent regardless of the acquisition environment. This specification formally tests whether the acquisition environment moderates the diagnostic group differences, serving to determine if the MDD effects are robust across experimental contexts.

| **Table S2.** Test of Group Differences Between Acquisition Contexts (**fMRI vs. Behavioral**) | | | | |
| --- | --- | --- | --- | --- |
| **Fixed Effect (Interaction Term)** | **Estimate** | **Std. Error** | **t value** | **p value** |
| ***b^+^*** x GroupMDD x ContextMRI | 0.109 | 0.1051 | 1.037 | 0.301 |
| ***b^-^*** x GroupMDD x ContextMRI | 0.2309 | 0.1125 | 2.053 | **0.042 *** |
| ***r^+^*** x GroupMDD x ContextMRI | -0.0606 | 0.1601 | -0.379 | 0.705 |
| ***r^-^*** x GroupMDD x ContextMRI | -0.4303 | 0.3235 | -1.33 | 0.185 |
| ***b^+^r^+^*** x GroupMDD x ContextMRI | -0.5604 | 0.293 | -1.913 | 0.056 |
| ***b^+^r^-^*** x GroupMDD x ContextMRI | 0.546 | 0.5076 | 1.075 | 0.282 |
| ***b^-^r^+^*** x GroupMDD x ContextMRI | -1.383 | 0.7611 | -1.818 | 0.071 |
| ***b^-^r^-^*** x GroupMDD x ContextMRI | 0.5233 | 0.9072 | 0.577 | 0.565 |

The analysis confirmed that the core MDD-specific behavioral patterns were consistent across acquisition settings (Table S2). Specifically, the three-way interactions for invest-loss (beta = 0.546, p = .282) and short-gain (beta = 0.523, p = .565) were non-significant, indicating that the group difference in responses to outcomes following negative market returns did not differ between acquisition contexts. While a significant three-way interaction was found for the influence of past negative bets on subsequent bet amount (beta = 0.231, p = .042), the overall stability of the outcome-based interactions supported the generalizability of the main clinical results.

**Supplementary Analysis 2: Unmedicated-MDD vs. Healthy Control - Matched Analysis**

To confirm the observed behavioral divergence was independent of pharmacological treatment, a sensitivity analysis compared the unmedicated MDD subsample (N=86) against the Healthy Control group (N=72). This analysis utilized the same iterative ensemble matching procedure (1:1 optimal matching on Age, Race, and Education, with exact match on Race, followed by 15,000 iterations of subsampling and linear mixed-effects modeling).

The behavioral divergence in MDD in response to outcomes following negative market returns remained highly significant (Table S3): invest-loss x Group (beta = -0.727, p < 0.001) and short-gain x Group (beta = -0.885, p < 0.001). Furthermore, a significant Short-Loss x Group divergence also emerged in this unmedicated comparison (beta = 0.356, p = 0.003).

**Supplementary Analysis 3: Medication Status-related Differences**

The percentage of medication-free participants differed between the total MDD cohort and the nested fMRI subsample (Total: 19.2% medicated vs. fMRI: 51.9% medicated; chi-square, p < .001; see Table 1). Therefore, to determine whether psychotropic medication use explained heterogeneity in response to outcome variables within the MDD participants, we compared a matched subset of all medicated (N=20) MDD subjects with matched (on age and education) unmedicated (N=20) MDD participants.

In this matched analysis, medication status did not significantly moderate the two key outcome effects under negative contextual valence that distinguished MDD from healthy controls in the main analysis, namely invest-loss (***b^+^r-*** × medicated: β = -0.03, p = .950) and short-gain (***b^-^r^-^*** × medicated: β = 1.25, p = .177; Table S3). There was only a trend-level interaction for invest-gain outcomes (***b^+^r^+^*** × medicated: β = -0.54, p = .059). No other medication-related interactions were significant.

| **Table S3. Supplementary Analyses Across Medication and Diagnostic Status** | | | | |
| --- | --- | --- | --- | --- |
| **Effect** | **Supplementary Analyses Unmedicated MDD vs. HC** | | | |
|  | **Beta** | **CI_low** | **CI_up** | **p** |
| **(Intercept)** | 0.078 | 0.066 | 0.088 | <.001*** |
| ***b^+^*** | 0.309 | 0.28 | 0.339 | <.001*** |
| ***b^-^*** | -0.354 | -0.376 | -0.33 | <.001*** |
| ***r^+^*** | 0.045 | 0.025 | 0.082 | 0.001*** |
| ***r-*** | -0.844 | -0.939 | -0.768 | <.001*** |
| ***b^+^r^+^*** | -0.484 | -0.62 | -0.383 | <.001*** |
| ***b^+^r-*** | 0.049 | -0.19 | 0.32 | 0.482 |
| ***b^-^r^+^*** | 0.729 | 0.529 | 0.928 | <.001*** |
| ***b^-^r^-^*** | 1.009 | 0.681 | 1.249 | <.001*** |
| **Age** | -0.001 | -0.001 | 0 | 0.001*** |
| **Race** | -0.007 | -0.014 | 0 | 0.056 |
| **Education** | 0.009 | -0.001 | 0.019 | 0.073 |
| **Group** | 0.004 | -0.004 | 0.011 | 0.312 |
| **b+:Group** | -0.029 | -0.062 | 0.005 | 0.087 |
| **b-:Group** | 0.026 | -0.005 | 0.058 | 0.097 |
| **r+:Group** | 0.085 | 0.042 | 0.129 | <.001*** |
| **r-:Group** | 0.126 | 0.027 | 0.233 | 0.011* |
| **b+r+:Group** | 0.088 | -0.072 | 0.245 | 0.263 |
| **b+r-:Group** | -0.727 | -1.018 | -0.451 | <.001*** |
| **b-r+:Group** | 0.356 | 0.117 | 0.596 | 0.003** |
| **b-r-:Group** | -0.885 | -1.181 | -0.519 | <.001*** |
| **Note:** Supplementary analyses examining the effect of medication status on model results. Results are from the same linear mixed-effects model excluding medicated MDD subjects with iterative ensemble matching procedure (10,000 iterations) comparing the unmedicated MDD subsample (N=86) to Healthy Controls (N=72), reporting the mean of the resulting empirical distribution. *p*-values and 95% Confidence Intervals (CI) are derived from the empirical distribution of coefficients. Healthy Control (HC) group served as the reference category. Significance levels: **p* < 0.05, ***p* < 0.01, ****p* < 0.001. | | | | |

**Table S4 Cluster-corrected whole-brain activation results by contrast and group**

| **Model** | **Contrast** | **Group** | **Peak region** | **Cluster size**  **(voxels)** | **Cluster p** | **Peak Z** | **MNI x** | **MNI y** | **MNI z** |
| --- | --- | --- | --- | --- | --- | --- | --- | --- | --- |
| Market return (r) | r^-^ | Across | Right Superior Frontal Gyrus | 327 | 1.19E-07 | 5.65 | 10.5 | 17.5 | 57 |
|  |  |  | Right Insular Cortex | 290 | 5.36E-07 | 4.74 | 31.5 | 11.5 | -18 |
|  |  |  | Right Frontal Pole | 238 | 4.23E-06 | 4.32 | 13.5 | 59.5 | 9 |
|  |  |  | Left Frontal Orbital Cortex | 231 | 5.66E-06 | 4.54 | -43.5 | 17.5 | -6 |
|  |  |  | Right Supramarginal Gyrus | 152 | 0.00018 | 4.84 | 52.5 | -42.5 | 15 |
|  |  |  | Right Cingulate Gyrus | 137 | 0.00037 | 4.49 | 7.5 | 20.5 | 33 |
|  |  |  | Left Frontal Pole | 98 | 0.00276 | 4.83 | -25.5 | 50.5 | 30 |
|  |  |  | Right Precuneous Cortex | 70 | 0.0138 | 4.17 | 16.5 | -63.5 | 27 |
|  |  |  | Right Cingulate Gyrus | 57 | 0.031 | 4.75 | 1.5 | -18.5 | 39 |
|  |  | HC | Right Superior Frontal Gyrus | 111 | 0.00135 | 4.59 | 10.5 | 17.5 | 57 |
|  |  |  | Right Frontal Operculum Cortex | 104 | 0.00195 | 3.77 | 46.5 | 17.5 | -3 |
|  |  |  | Right Frontal Pole | 79 | 0.00792 | 3.91 | 22.5 | 47.5 | 18 |
|  |  | MDD | Left Insular Cortex | 184 | 4.05E-05 | 4.75 | -40.5 | 8.5 | -6 |
|  |  |  | Right Superior Frontal Gyrus | 162 | 0.00011 | 4.38 | 7.5 | 17.5 | 63 |
|  |  |  | Right Paracingulate Gyrus | 82 | 0.00665 | 4.05 | 10.5 | 53.5 | 0 |
|  |  |  | Right Cingulate Gyrus | 58 | 0.0287 | 3.82 | 10.5 | 17.5 | 27 |
|  |  |  | Left Frontal Pole | 54 | 0.0372 | 4.09 | -25.5 | 47.5 | 24 |
|  | r^+^ | Across | Right Occipital Fusiform Gyrus | 425 | 5.89E-09 | 5.18 | 25.5 | -69.5 | -6 |
|  |  |  | Right Frontal Operculum Cortex | 367 | 5.96E-08 | 4.81 | 43.5 | 20.5 | -3 |
|  |  |  | Right Superior Frontal Gyrus | 183 | 5.03E-05 | 4.07 | 25.5 | 8.5 | 45 |
|  |  |  | Left Middle Frontal Gyrus | 157 | 0.000162 | 4.85 | -31.5 | 2.5 | 48 |
|  |  |  | Left Insular Cortex | 106 | 0.00196 | 5.09 | -31.5 | 14.5 | -6 |
|  |  |  | Left Lateral Occipital Cortex | 100 | 0.0027 | 4.59 | -25.5 | -78.5 | 21 |
|  |  |  | Right Caudate | 87 | 0.00549 | 4.99 | 10.5 | 8.5 | 0 |
|  |  |  | Right Brain-Stem | 56 | 0.0348 | 5.42 | 4.5 | -30.5 | -3 |
|  |  | HC | Left Middle Frontal Gyrus | 93 | 0.00397 | 4.57 | -34.5 | 2.5 | 48 |
|  |  |  | Right Insular Cortex | 67 | 0.0177 | 3.91 | 31.5 | 20.5 | 0 |
|  |  |  | Right Caudate | 62 | 0.024 | 4.81 | 10.5 | 5.5 | 0 |
|  |  | MDD | Right Intracalcarine Cortex | 257 | 2.44E-06 | 4.72 | 7.5 | -75.5 | 9 |
|  |  |  | Right Frontal Orbital Cortex | 111 | 0.00153 | 4.44 | 37.5 | 26.5 | -9 |
| Fictive error (\|r\| - br) | b^-^r^-^ | Across | Right Frontal Orbital Cortex | 1943 | 1.45E-24 | 5.77 | 28.5 | 17.5 | -18 |
|  |  |  | Right Angular Gyrus | 608 | 3.24E-11 | 5.36 | 46.5 | -57.5 | 36 |
|  |  |  | Left Inferior Frontal Gyrus | 419 | 1.05E-08 | 5.28 | -52.5 | 23.5 | 6 |
|  |  |  | Right Precuneous Cortex | 239 | 6.32E-06 | 4.47 | 16.5 | -60.5 | 30 |
|  |  |  | Left Frontal Pole | 86 | 0.00668 | 4.54 | -19.5 | 50.5 | 24 |
|  |  |  | Left Middle Frontal Gyrus | 64 | 0.0237 | 4.37 | -37.5 | 14.5 | 36 |
|  |  |  | Left Angular Gyrus | 61 | 0.0285 | 3.82 | -52.5 | -57.5 | 33 |
|  |  | HC | Right Frontal Orbital Cortex | 308 | 4.77E-07 | 5.07 | 28.5 | 17.5 | -18 |
|  |  |  | Right Superior Frontal Gyrus | 162 | 0.000154 | 4.1 | 10.5 | 17.5 | 63 |
|  |  |  | Right Lateral Occipital Cortex | 126 | 0.000829 | 4 | 43.5 | -69.5 | 36 |
|  |  |  | Right Frontal Pole | 75 | 0.0122 | 3.97 | 28.5 | 53.5 | -3 |
|  |  | MDD | Left Cingulate Gyrus | 430 | 6.94E-09 | 4.34 | -10.5 | 35.5 | 9 |
|  |  |  | Left Insular Cortex | 359 | 5.96E-08 | 5.3 | -43.5 | 14.5 | -6 |
|  |  |  | Right Frontal Orbital Cortex | 202 | 2.73E-05 | 5.09 | 37.5 | 26.5 | -6 |
|  |  |  | Right Angular Gyrus | 100 | 0.00309 | 4.27 | 46.5 | -57.5 | 36 |
|  | b^+^r^-^ | Across | Right Superior Frontal Gyrus | 709 | 7.06E-14 | 5.73 | 10.5 | 11.5 | 60 |
|  |  |  | Right Insular Cortex | 95 | 0.00196 | 4.3 | 37.5 | 11.5 | -3 |
|  |  |  | Right Supramarginal Gyrus | 89 | 0.00282 | 4.01 | 58.5 | -42.5 | 12 |
|  |  |  | Left Insular Cortex | 84 | 0.00384 | 4.61 | -40.5 | 11.5 | 0 |
|  |  |  | Left Precuneous Cortex | 60 | 0.0185 | 4.57 | -10.5 | -54.5 | 60 |
|  |  |  | Left Frontal Pole | 57 | 0.0228 | 4.57 | -37.5 | 44.5 | 33 |
|  |  |  | Right Occipital Fusiform Gyrus | 52 | 0.0325 | 3.91 | 25.5 | -63.5 | -18 |
|  |  | HC | Right Superior Frontal Gyrus | 76 | 0.00633 | 4.87 | 10.5 | 11.5 | 60 |
|  |  | MDD | Right Superior Frontal Gyrus | 85 | 0.00359 | 3.98 | 13.5 | 11.5 | 60 |
|  |  |  | Right Cingulate Gyrus | 67 | 0.0114 | 3.81 | 7.5 | 14.5 | 33 |
|  | b^+^r^+^ | Across | Right Paracingulate Gyrus | 888 | 3.34E-15 | 5.06 | 7.5 | 32.5 | 27 |
|  |  |  | Right Insular Cortex | 376 | 1.79E-08 | 5.51 | 34.5 | 14.5 | -9 |
|  |  |  | Left Superior Parietal Lobule | 258 | 1.55E-06 | 4.48 | -34.5 | -51.5 | 48 |
|  |  |  | Left Insular Cortex | 184 | 3.56E-05 | 5.92 | -34.5 | 17.5 | -3 |
|  |  |  | Left Middle Frontal Gyrus | 80 | 0.00714 | 4.14 | -40.5 | 26.5 | 42 |
|  |  |  | Right Precuneous Cortex | 63 | 0.0203 | 4.57 | 10.5 | -51.5 | 48 |
|  |  |  | Ventral Tegmental Area (VTA) | 58 | 0.028 | 4.34 | -4.5 | -15.5 | -12 |
|  |  |  | Right Caudate | 53 | 0.039 | 4.53 | 10.5 | 8.5 | 0 |
|  |  | HC | Right Paracingulate Gyrus | 281 | 6.56E-07 | 4.64 | 4.5 | 41.5 | 30 |
|  |  |  | Right Middle Frontal Gyrus | 216 | 8.88E-06 | 4.22 | 55.5 | 11.5 | 39 |
|  |  |  | Right Insular Cortex | 166 | 8.24E-05 | 4.28 | 31.5 | 23.5 | 0 |
|  |  |  | Left Middle Frontal Gyrus | 117 | 0.000921 | 3.93 | -34.5 | 2.5 | 57 |
|  |  |  | Left Insular Cortex | 86 | 0.00506 | 4.97 | -37.5 | 17.5 | -6 |
|  |  |  | Right Caudate | 82 | 0.00638 | 4.41 | 10.5 | 5.5 | 0 |
|  |  | MDD | Left Supramarginal Gyrus | 134 | 0.000386 | 4.59 | -61.5 | -30.5 | 36 |
|  |  |  | Right Insular Cortex | 100 | 0.00229 | 4.8 | 34.5 | 14.5 | -9 |
|  |  |  | Left Superior Frontal Gyrus | 74 | 0.0103 | 4.06 | -19.5 | -0.5 | 54 |
|  |  |  | Left Insular Cortex | 52 | 0.0419 | 4.29 | -31.5 | 17.5 | -3 |
| **Note.** Results are cluster-corrected at α < .05 using a cluster-forming threshold of p < .001. Cluster size is reported in voxels. Cluster p values are FSL cluster-corrected p values. Peak Z indicates the maximum Z-statistic within each cluster, and MNI x, y, and z indicate the coordinates of that peak voxel. Peak regions were assigned using the Harvard-Oxford Cortical and Subcortical Structural Atlases at the peak MNI coordinate, with manual relabeling of midbrain/brainstem peaks where appropriate. Labels are approximate and clusters may extend beyond the listed peak region. No suprathreshold clusters were observed for the short-loss (b⁻r⁺) contrast or for whole-brain group-difference contrasts | | | | | | | | | |

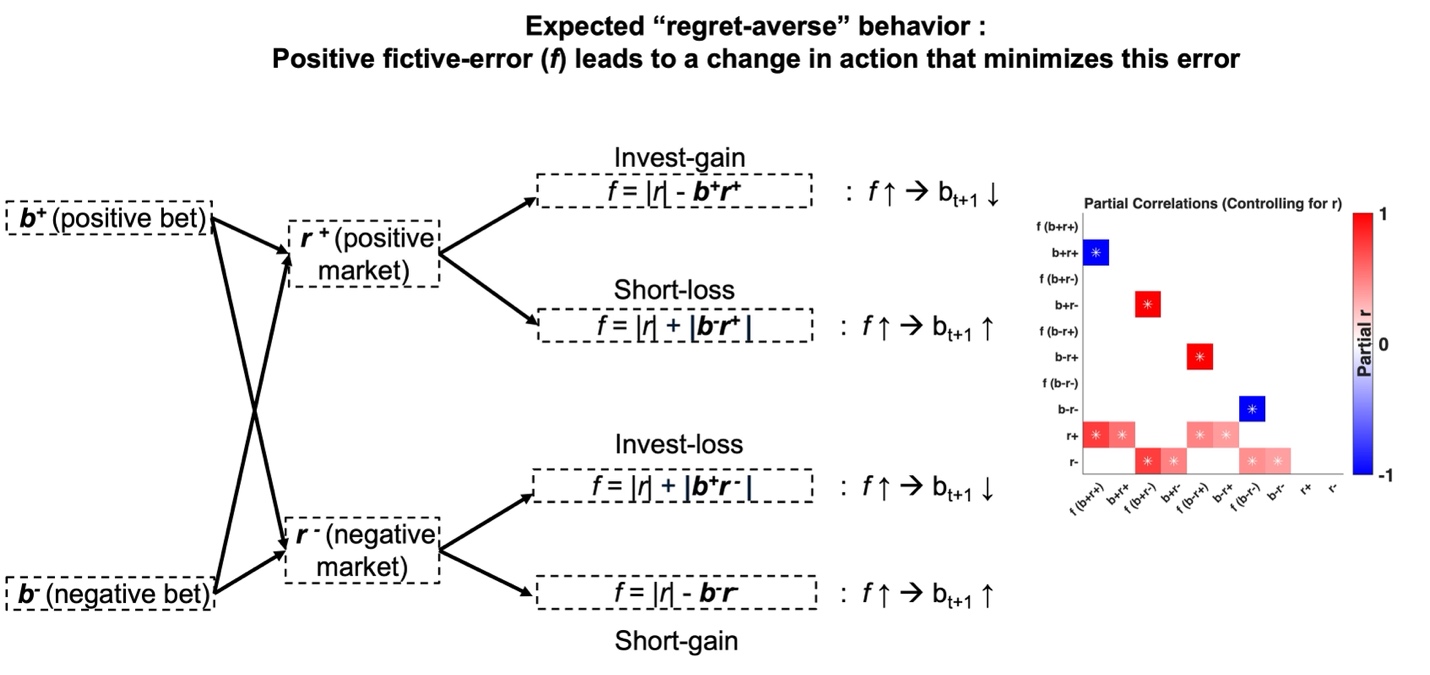

**Figure S1.** The schematic illustrates how fictive error signals—computed as the difference between actual and best-possible outcomes—are theorized to guide subsequent investment behavior. For each combination of bet direction and market change, a positive fictive error (i.e., high regret) is expected to prompt a change in future bets that would reduce regret. In gain scenarios (b⁺r⁺, b⁻r⁻), smaller gains reflect higher regret, which is expected to increase the tendency to adjust future bets in a regret-minimizing direction. For example, following an “invest-gain” (b⁺r⁺) outcome, a small gain after a low bet would imply high fictive error (i.e., regret), predicting that the individual will increase their bet in the next trial to rectify the regret-inducing behavior (i.e., invest-gain and next bets are inversely correlated). In loss scenarios (b⁺r⁻, b⁻r⁺), larger losses reflect higher fictive error, similarly predicting greater adjustment in future behavior. For instance, following a “short-loss” (b⁻r⁺) outcome, a large loss after a high short bet would imply high regret, and the model predicts that individuals will reduce the magnitude of short betting on the subsequent trial (i.e., short-loss and next bets are positively correlated). The partial correlation matrix illustrates the relationships between actual outcome and fictive errors from the actual data while controlling for market returns (r⁺ or r⁻), isolating the associations from market-driven covariance. White asterisks indicate correlations with absolute values ≥ 0.3. For fMRI analysis, neutral trials (b=0) were classified into the "gain" regressors (invest-gain and short-gain) to specifically investigate the neural correlates of increasing fictive error. Within the gain domain, a zero bet represents the maximum possible fictive error (regret). In contrast, for loss outcomes, a zero bet represents the minimum possible fictive error (relief), signaling successful loss avoidance.

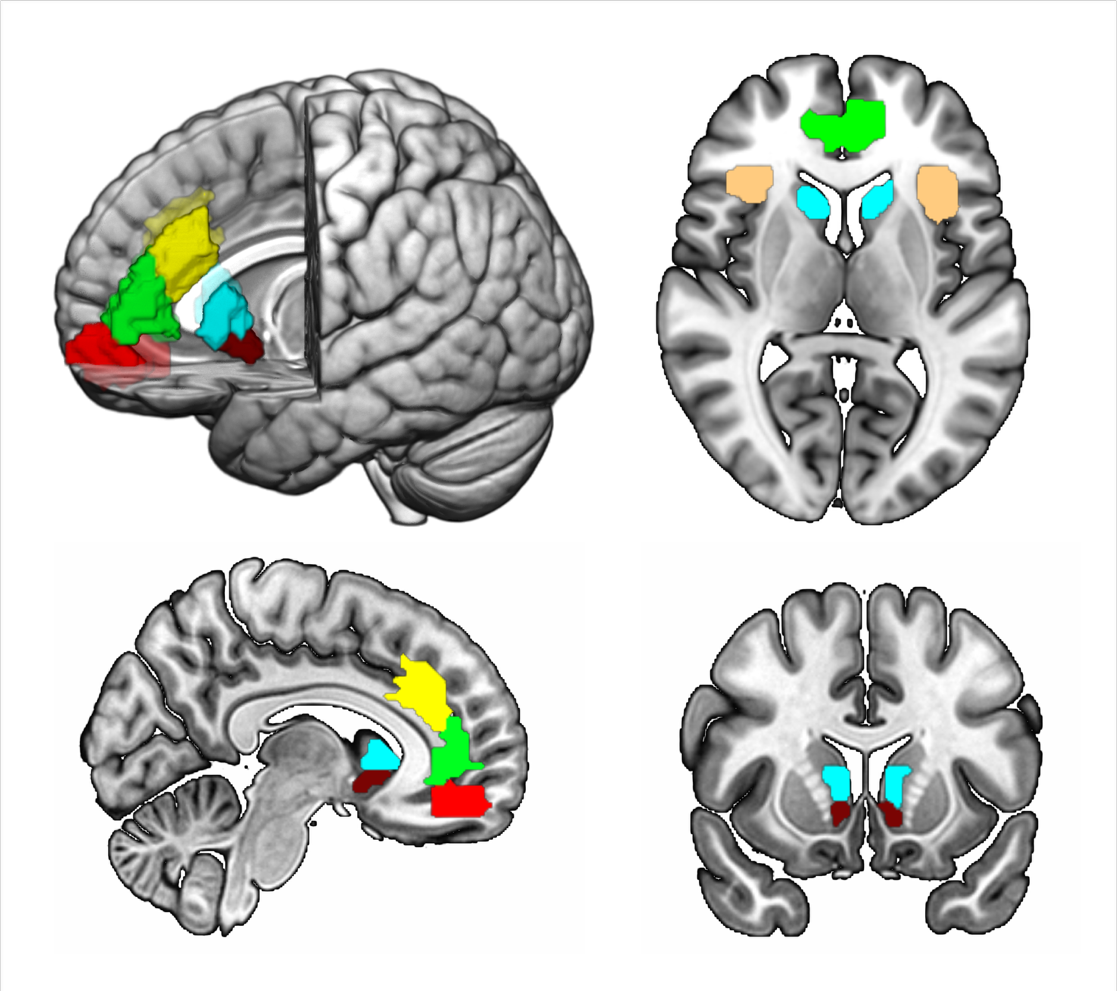

**Figure S2.** Placement of regions of interest (ROIs) used in the study. The ventral caudate (vCaud; lightblue) and nucleus accumbens (NAcc = maroon) were defined using the Harvard-Oxford subcortical atlas in FSL, with vCaud restricted to voxels inferior to z = 7 of the caudate. The dorsal and rostral anterior cingulate cortex (dACC = yellow; rACC=green), anterior insula (aIns=chardonnay) and ventromedial prefrontal cortex/medial orbitofrontal cortex (vmPFC/mOFC = red) were defined using function-connectivity-based Schaefer 2018 200-parcellation atlas. ROIs are overlaid on a standard MNI template for visualization.

***Group differences in market return* (*r_t_*) *related brain activity***

Across participants, positive market returns elicited significant clusters of activation in the ventral striatum/caudate, bilateral insula/frontal operculum, and frontal cortical regions, including superior and middle frontal gyri (Table S4). Group-specific analyses showed HC participants had suprathreshold clusters in ventral striatum and anterior insula (Figure S3A, top row), while MDD participants showed suprathreshold activity only in anterior insula (Figure S3A, bottom row). In response to negative market returns, significant activation was observed in the anterior insula and mid-frontal regions across all participants (Table S4). Notably, significant activation in the anterior cingulate was observed exclusively within the MDD group (Figure S3B, bottom row). Whole-brain group comparisons revealed no significant differences in activation patterns for either positive or negative market returns.

ROI analyses demonstrated a dissociation (Figure S3C-D): HC participants showed stronger ventral caudate responses to positive market returns compared with MDD participants (*p* = 0.009), while MDD participants exhibited greater vmPFC, dACC, and rACC activation to negative market returns (all uncorrected *p*s < .05), with only the ACC effects surviving FDR correction.

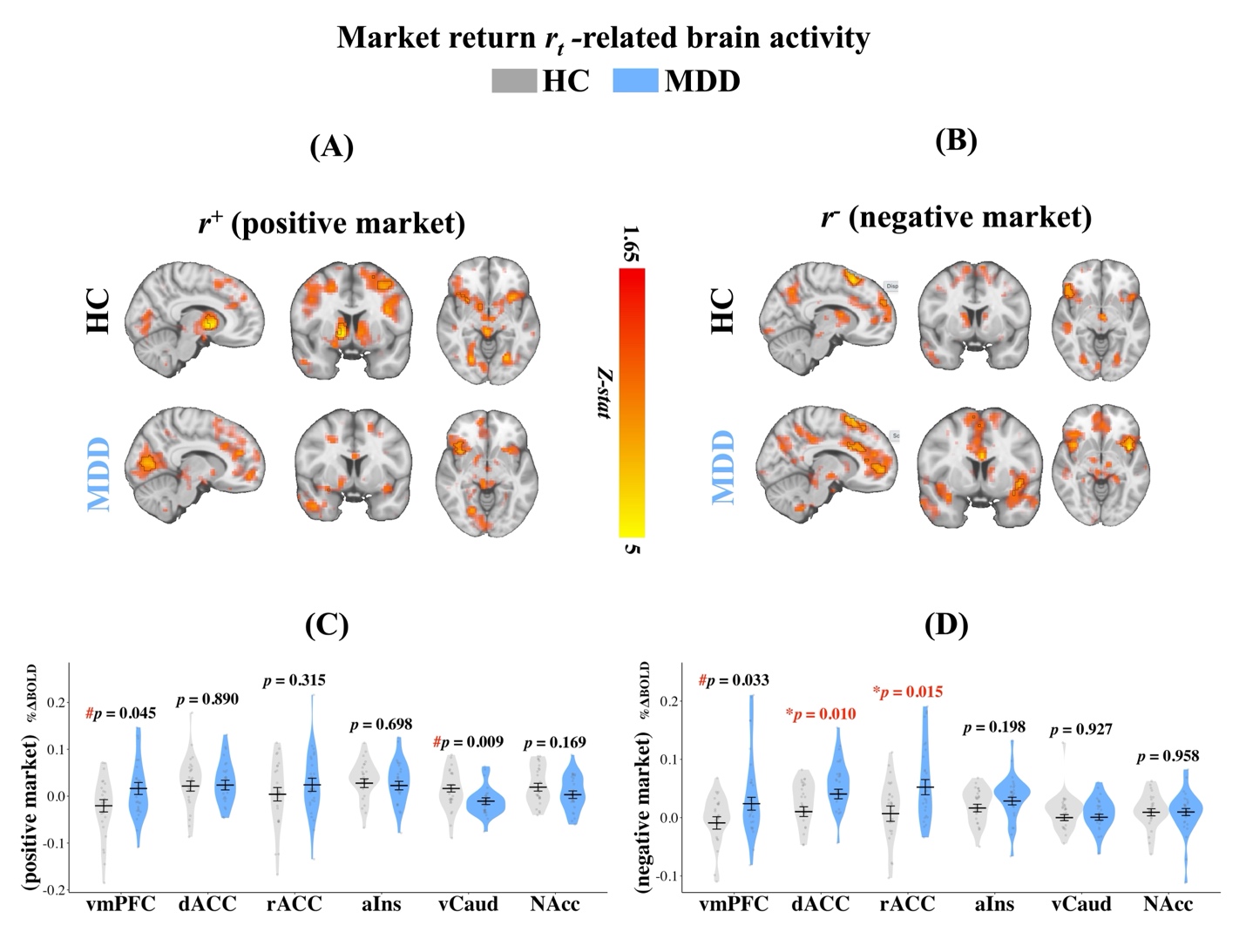
**Figure S3. Depression is associated with altered processing of market returns. (A–B)** Whole-brain activation maps showing market return-related activity (|rt|) for **(A) positive** and **(B) negative market returns.** Separate rows show HC and MDD participants. Opaque regions outlined in black indicate suprathreshold clusters surviving cluster correction at α < .05, using a cluster-forming threshold of p < .001. Faint overlays show subthreshold positive Z-statistics above Z = 1.645 (~p < .05) for visualization only. Warm colors indicate positive Z-statistics on a common display scale. **(C–D) Region-of-interest (ROI) analysis** quantifying group differences for **(C) positive** and **(D) negative market return**-related changes in BOLD signal. Dots represent covariate-adjusted % change in BOLD signal (controlling for age and sex); violin plots show density; error bars indicate mean +/- SEM. Extreme outliers were excluded using a 3.5 × IQR threshold (maximum one subject excluded for displayed ROIs); sensitivity analyses confirmed results were unchanged by outlier inclusion and remained unmoderated by MDD medication status. Statistical significance was assessed with Type III ANOVA, with FDR correction applied across the six evaluated ROI targets. Asterisks denote effects surviving FDR correction across the six tested ROIs (* p < 0.05, ** p < 0.01); a red pound sign (#) denotes uncorrected p < 0.05 that did not survive FDR correction. Abbreviations: vmPFC, ventromedial Prefrontal Cortex; dACC, dorsal Anterior Cingulate Cortex; rACC, rostral Anterior Cingulate Cortex; aIns, anterior Insula; vCaud, ventral Caudate; NAcc, Nucleus Accumbens; HC, Healthy Controls; MDD, Major Depressive Disorder.

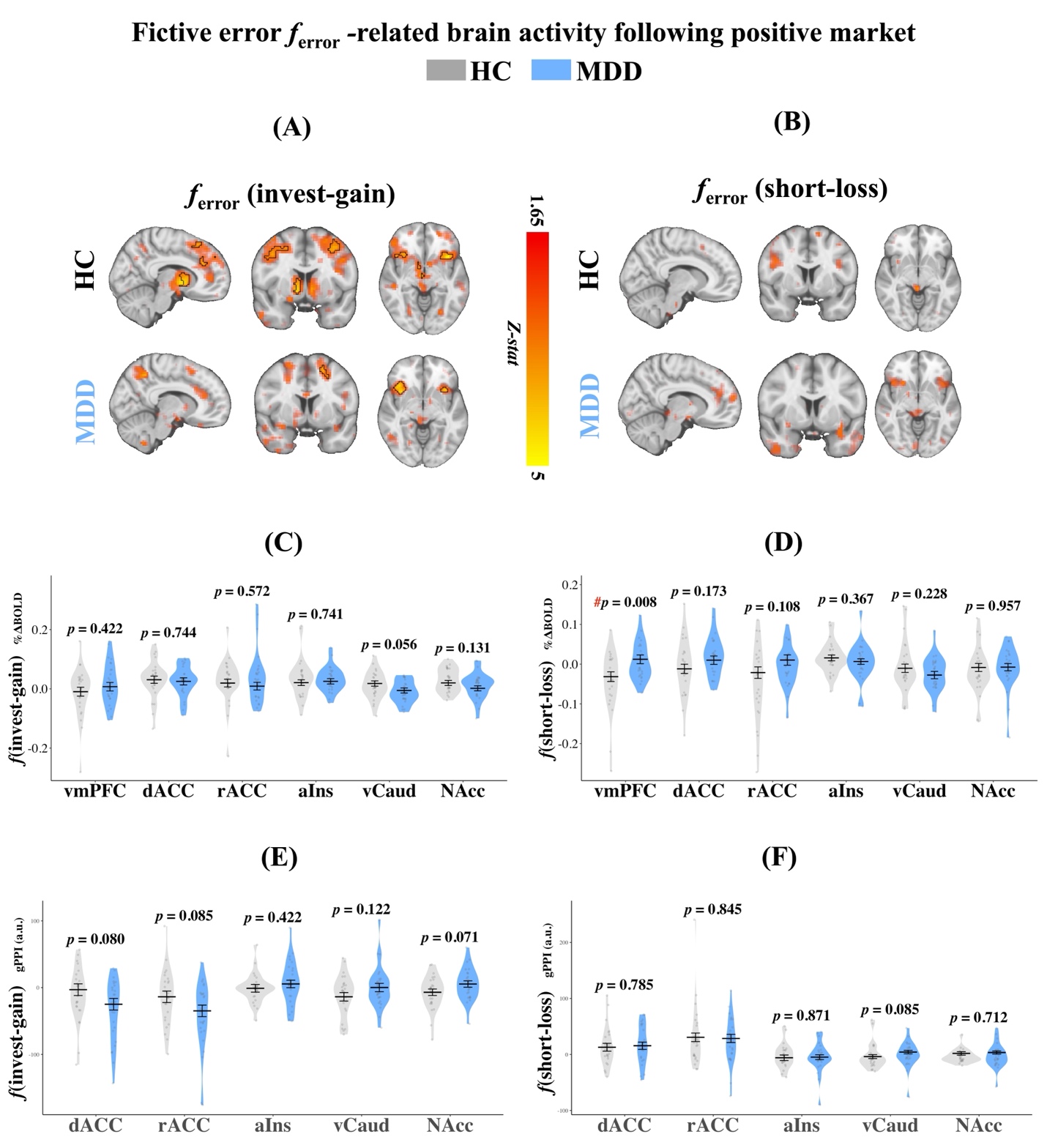

**Figure S4. Group differences in fictive-error processing following positive market returns at the activation and functional-connectivity levels.** **(A) Invest-gain and (B) short-loss fictive errors**, shown separately for HC and MDD participants. Opaque regions outlined in black indicate suprathreshold clusters surviving cluster correction at α < .05, using a cluster-forming threshold of p < .001. Faint overlays show subthreshold positive Z-statistics above Z = 1.645 (~p < .05) for visualization only. Warm colors indicate positive Z-statistics on a common display scale. **(C–D) ROI analyses of activation differences** for (**C) invest-gain and (D) short-loss fictive errors**. Points represent covariate-adjusted % change in BOLD signal from robust linear regression models controlling for age and sex and weighted by trial count and entropy; violin plots show the distribution and error bars indicate mean ± SEM. No group differences were observed for invest-gain errors. For short-loss errors, MDD participants showed greater vmPFC activity than HCs at the uncorrected level (p_uncorr_ = 0.008), but this effect did not survive FDR correction. No significant group differences were observed in vCaud or NAcc for either error type. Results were unchanged in sensitivity analyses excluding extreme outliers (>3.5 × IQR; maximum 2 exclusions per ROI) and were not moderated by medication status (all ps > .05). Asterisks denote effects surviving FDR correction across the six tested ROIs (* p < 0.05, ** p < 0.01); a red pound sign (#) denotes uncorrected p < 0.05 that did not survive FDR correction. (E–F) ROI analyses of task-dependent functional connectivity (gPPI) for (E) invest-gain and (F) short-loss fictive errors. Points represent covariate-adjusted gPPI estimates (a.u.), violin plots show the distribution, and horizontal lines with error bars indicate estimated marginal means ± SEM. No significant group differences in vmPFC/mOFC coupling were observed for either error type. Extreme outliers were excluded using a 3.5 × IQR threshold (maximum one subject excluded for displayed ROIs). Statistical significance was assessed with Type III ANOVA, with FDR correction applied across the five evaluated ROI targets. Abbreviations: vmPFC, ventromedial Prefrontal Cortex; dACC, dorsal Anterior Cingulate Cortex; rACC, rostral Anterior Cingulate Cortex; aIns, anterior Insula; vCaud, ventral Caudate; NAcc, Nucleus Accumbens; HC, Healthy Controls; MDD, Major Depressive Disorder.

**Table S5. ROI activity–symptom associations within the MDD group**

| **Context** | **Contrast** | **ROI** | **Symptom** | **β** | **95% CI** | **p** | **pFDR** | **Sig** |
| --- | --- | --- | --- | --- | --- | --- | --- | --- |
| **Negative market** | **Invest-loss (b+r-)** | **vmPFC** | Depression | 0.036 | [0.01, 0.062] | 0.009 | 0.034 | ***** |
|  |  |  | Anhedonia (SHAPS) | 0.014 | [-0.016, 0.045] | 0.332 | 0.454 |  |
|  |  |  | TEPS-A | -0.009 | [-0.033, 0.015] | 0.454 | 0.454 |  |
|  |  |  | TEPS-C | -0.009 | [-0.032, 0.015] | 0.449 | 0.454 |  |
|  |  |  | Anxiety | 0.019 | [-0.011, 0.05] | 0.204 | — |  |
|  |  | **dACC** | Depression | 0.007 | [-0.015, 0.03] | 0.502 | 0.812 |  |
|  |  |  | Anhedonia (SHAPS) | -0.004 | [-0.029, 0.021] | 0.734 | 0.812 |  |
|  |  |  | TEPS-A | 0.002 | [-0.019, 0.024] | 0.812 | 0.812 |  |
|  |  |  | TEPS-C | 0.003 | [-0.018, 0.025] | 0.757 | 0.812 |  |
|  |  |  | Anxiety | -0.004 | [-0.029, 0.02] | 0.720 | — |  |
|  |  | **rACC** | Depression | 0.032 | [0.005, 0.06] | 0.024 | 0.097 | **#** |
|  |  |  | Anhedonia (SHAPS) | 0.021 | [-0.013, 0.056] | 0.211 | 0.379 |  |
|  |  |  | TEPS-A | -0.011 | [-0.043, 0.02] | 0.457 | 0.457 |  |
|  |  |  | TEPS-C | -0.017 | [-0.048, 0.015] | 0.284 | 0.379 |  |
|  |  |  | Anxiety | 0.020 | [-0.014, 0.055] | 0.232 | — |  |
|  |  | **aIns** | Depression | -0.009 | [-0.028, 0.009] | 0.291 | — |  |
|  |  |  | Anhedonia (SHAPS) | 0.005 | [-0.01, 0.021] | 0.477 | — |  |
|  |  |  | TEPS-A | 0.000 | [-0.017, 0.016] | 0.974 | — |  |
|  |  |  | TEPS-C | 0.000 | [-0.017, 0.017] | 0.996 | — |  |
|  |  |  | Anxiety | -0.015 | [-0.034, 0.004] | 0.116 | — |  |
|  | **Short-gain (b-r-)** | **vmPFC** | Depression | 0.030 | [0.001, 0.059] | 0.046 | 0.061 | **#** |
|  |  |  | Anhedonia (SHAPS) | 0.036 | [0.005, 0.067] | 0.024 | 0.048 | ***** |
|  |  |  | TEPS-A | -0.032 | [-0.06, -0.005] | 0.023 | 0.048 | ***** |
|  |  |  | TEPS-C | -0.020 | [-0.047, 0.008] | 0.157 | 0.157 |  |
|  |  |  | Anxiety | 0.013 | [-0.028, 0.054] | 0.510 | — |  |
|  |  | **dACC** | Depression | -0.005 | [-0.039, 0.029] | 0.751 | 0.751 |  |
|  |  |  | Anhedonia (SHAPS) | 0.023 | [-0.009, 0.054] | 0.150 | 0.300 |  |
|  |  |  | TEPS-A | -0.023 | [-0.053, 0.006] | 0.116 | 0.300 |  |
|  |  |  | TEPS-C | -0.012 | [-0.042, 0.018] | 0.412 | 0.550 |  |
|  |  |  | Anxiety | -0.023 | [-0.059, 0.013] | 0.205 | — |  |
|  |  | **rACC** | Depression | 0.011 | [-0.012, 0.035] | 0.332 | 0.336 |  |
|  |  |  | Anhedonia (SHAPS) | 0.025 | [0.002, 0.047] | 0.031 | 0.062 | **#** |
|  |  |  | TEPS-A | -0.020 | [-0.039, -0.002] | 0.031 | 0.062 | **#** |
|  |  |  | TEPS-C | -0.010 | [-0.03, 0.011] | 0.336 | 0.336 |  |
|  |  |  | Anxiety | -0.007 | [-0.037, 0.024] | 0.646 | — |  |
|  |  | **aIns** | Depression | 0.001 | [-0.028, 0.03] | 0.938 | — |  |
|  |  |  | Anhedonia (SHAPS) | -0.009 | [-0.035, 0.018] | 0.492 | — |  |
|  |  |  | TEPS-A | 0.008 | [-0.018, 0.033] | 0.528 | — |  |
|  |  |  | TEPS-C | 0.004 | [-0.02, 0.027] | 0.742 | — |  |
|  |  |  | Anxiety | 0.006 | [-0.028, 0.039] | 0.719 | — |  |

**Table S5.** Standardized beta coefficients, 95% confidence intervals (CI), and p-values for clinical associations with fictive error signals following negative market returns. Influential data points were excluded using a Cook’s Distance threshold of 4/N, where N represents the sample size for each specific model. **Abbreviations:** SHAPS = Snaith-Hamilton Pleasure Scale; TEPS-A = Temporal Experience of Pleasure Scale (Anticipatory); TEPS-C = Temporal Experience of Pleasure Scale (Consummatory). Depression severity and Anxiety scores represent standardized average scores across multiple scales; refer to the "Clinical symptom scores" section in the main text for the specific measures used. vCaud = ventral Caudate; vmPFC = ventromedial prefrontal cortex; dACC = dorsal anterior cingulate cortex; rACC = rostral anterior cingulate cortex. ****p < .001, *p < .05, • p < .10.*

**Clinical Score Comparison by Medication Status**

To assess whether medication status significantly impacted depression severity, independent-samples *t*-tests were conducted within the Major Depressive Disorder (MDD) cohort to compare clinical scores between medicated and unmedicated patients. The analysis, restricted to the MDD group, found no statistically significant differences (*p* > 0.05) in any of the tested clinical scores between medicated and unmedicated participants. Results are reported in Table S6.

| **Table S6. Comparison of Clinical Score Means Between Medicated and Unmedicated Patients with Major Depressive Disorder (MDD)** | | | | | |
| --- | --- | --- | --- | --- | --- |
| **Variable** | **N**  **(Med_Yes)** | **N**  **(Med_No)** | **Mean**  **(Med Yes)** | **Mean**  **(Med No)** | **P-value** |
| **SHAPS** | 19 | 91 | 33.1 | 32.1 | 0.723 |
| **TEPS (Anticipatory)** | 14 | 13 | 28.1 | 35 | 0.151 |
| **TEPS (Consummatory)** | 14 | 13 | 30.1 | 32.2 | 0.593 |
| **QIDS** | 19 | 91 | 13.5 | 13.5 | 0.999 |
| **Note:** N (Med Yes) and N (Med No) indicate the final sample size available for each measure after the exclusion of missing data. P-values were derived from independent-samples t-tests. Abbreviations: SHAPS, Snaith-Hamilton Pleasure Scale; TEPS, Temporal Experience of Pleasure Scale; QIDS, Quick Inventory of Depressive Symptomatology. | | | | | |
